# Faster bi-stable visual switching in psychosis

**DOI:** 10.1101/2023.02.13.23285774

**Authors:** Kyle W. Killebrew, Hannah R. Moser, Andrea N. Grant, Małgorzata Marjańska, Scott R. Sponheim, Michael-Paul Schallmo

## Abstract

Bi-stable stimuli evoke two distinct perceptual interpretations that alternate and compete for dominance. Bi-stable perception is thought to be driven at least in part by mutual suppression between distinct neural populations that represent each percept. Abnormal visual perception is observed among people with psychotic psychopathology (PwPP), and there is evidence to suggest that these visual deficits may depend on impaired neural suppression in visual cortex. However, it is not yet clear whether bi-stable visual perception is abnormal among PwPP. Here, we examined bi-stable perception in a visual structure-from-motion task using a rotating cylinder illusion in a group of 65 PwPP, 44 first-degree biological relatives, and 43 healthy controls. Data from a ‘real switch’ task, in which physical depth cues signaled real switches in rotation direction were used to exclude individuals who did not show adequate task performance. In addition, we measured concentrations of neurochemicals, including glutamate, glutamine, and γ-amino butyric acid (GABA), involved in excitatory and inhibitory neurotransmission. These neurochemicals were measured non-invasively in visual cortex using 7 tesla MR spectroscopy. We found that PwPP and their relatives showed faster bi-stable switch rates than healthy controls. Faster switch rates also correlated with significantly higher psychiatric symptom levels across all participants. However, we did not observe any significant relationships across individuals between neurochemical concentrations and SFM switch rates. Our results are consistent with a reduction in suppressive neural processes during structure-from-motion perception in PwPP, and suggest that genetic liability for psychosis is associated with disrupted bi-stable perception.

## 1. Introduction

Psychotic symptoms, which include hallucinations and delusions, are a defining feature of schizophrenia, schizoaffective disorder, and bipolar disorder. These symptoms can have a dramatic impact on a person’s ability to perform daily tasks (“Diagnostic and Statistical Manual of Mental Disorders,” 2013; Revheim et al., 2006). In addition to frank hallucinations, distorted perceptions of real stimuli are often reported, most often in the auditory domain, but with visual distortions being common as well (Thomas et al., 2007; Waters et al., 2014). This type of visual dysfunction can be studied using well-established and tested behavioral paradigms that allow the measurement of more subtle behavioral and cognitive disruptions (Waters et al., 2014). Studying visual dysfunction in people with psychotic psychopathology (PwPP) using behavioral paradigms may thus provide insight into the neural basis of psychotic symptoms, and may also inform a better understanding of basic sensory processing (Notredame et al., 2014; Phillips & Silverstein, 2013; Yoon et al., 2013).

Bi-stable perception is a phenomenon that occurs when the same external stimulus evokes two alternating dominant percepts. Since the early 20^th^ century (George, 1936; McDougall, 1906), researchers have studied these bi-stable stimuli by measuring the rate at which participants’ subjective percept switches from one interpretation to the other. There are many examples of this phenomenon, including both static and dynamic stimuli. Some static versions include the Necker cube, Rubin’s face-vase illusion, and binocular rivalry, whereas dynamic versions include the spinning dancer, point light walker, and the rolling wheel (Leopold & Logothetis, 1999; Martínez & Parra, 2018; Sterzer et al., 2009).

Another dynamic bi-stable percept is the rotating cylinder (Siegel & Andersen, 1990; Treue et al., 1991; Ullman, 1979). In this illusion, dots traverse from left to right and right to left in a rectangular area, speeding up when approaching the middle and slowing down when approaching the edge, which gives the impression that the dots are ‘painted’ on the surface of a translucent rotating cylinder. The rotating cylinder is an example of structure-from-motion (SFM) perception, which involves integrating many moving elements into one coherent percept, where the perceived object is defined by the movement of the elements (Kourtzi et al., 2008). This stimulus provides an opportunity to study the interaction between the perception of object motion and form.

The rotating cylinder is thought to involve neural processing at multiple levels within the visual hierarchy, including within regions selective for motion and shape processing. Functional magnetic resonance imagining (fMRI) studies have shown that responses in human medial temporal complex (hMT+) are selective for stimulus features including motion direction and coherence (Braddick et al., 2000; Rees et al., 2000; Smith et al., 2006). Additionally, hMT+ shows selectivity for motion in depth and structure-from-motion (Brouwer & van Ee, 2007; Freeman et al., 2012), suggesting that hMT+ may play an important role in the perception of the rotating cylinder illusion. Likewise, the lateral occipital complex (LOC), which plays an important role in object and form processing (Larsson & Heeger, 2006), has been shown to respond more strongly to rotation-in-depth SFM stimuli (versus scrambled or translational motion), indicating a role for LOC in the processing of motion-defined shapes (Freeman et al., 2012; Murray et al., 2002).

Abnormalities in laboratory assessments of visual perception have been reported in psychotic disorders, ranging from low to higher-level processes (Butler et al., 2008; Notredame et al., 2014; Phillips & Silverstein, 2013; Yoon et al., 2013), including both atypical motion perception and visual integration. For example, studies among PwPP have shown impairments in some aspects of visual motion perception (Chen et al., 1999, 2003, 2005, 2006, 2008; Kaliuzhna et al., 2020; Tadin et al., 2006; Zeljic et al., 2021). In general, these findings suggest there may be deficits in global motion perception associated with psychosis, while local motion perception may be relatively intact (Bennett et al., 2016; Carter et al., 2017; Chen et al., 2003). Visual integration is a process by which visual information is combined to create the percept of a more complex whole (Loffler, 2008; Roelfsema & de Lange, 2016). Integration of visual forms also involves processing across multiple levels within the visual hierarchy, and includes visual functions such as center-surround suppression, contour integration, border detection, shape representation, and figure-ground segmentation, all of which have been found to be impaired among PwPP (Chen et al., 2008; Dakin et al., 2005; Foxe et al., 2005; Keane et al., 2012, 2016; Kozma-Wiebe et al., 2006; Pokorny et al., 2020; Robol et al., 2013; M. P. Schallmo et al., 2013, 2015; Silverstein et al., 2006, 2009, 2012, 2015; Spencer et al., 2003; Spencer & Ghorashi, 2014; Tadin et al., 2006; Tibber et al., 2013; Wynn et al., 2015; Yang et al., 2013; Yeap et al., 2006). Thus, impairments in visual integration and visual motion perception may both be relevant for SFM perception in PwPP.

One potential explanation for abnormal vision in psychosis is a disruption in excitatory / inhibitory (E/I) balance (Foss-Feig et al., 2017; Lisman, 2012). This has been proposed in PwPP based on evidence showing disruption in glutamate (Javitt, 2004; Moghaddam & Javitt, 2012) and γ-amino butyric acid (GABA) signaling (Gonzalez-Burgos et al., 2010; Volk & Lewis, 2005). Bi-stable visual paradigms may provide a convenient behavioral marker of E/I balance in visual cortex. Specifically, the rate at which bi-stable stimuli alternate, or the duration for which one percept dominates, has been linked to excitatory (i.e., glutamatergic) and inhibitory (i.e., GABA-ergic) neural processes in humans (Ip et al., 2021; Mentch et al., 2019; Robertson et al., 2016; Sandberg et al., 2016; van Loon et al., 2013). If PwPP show different bi-stable switch rates versus controls, this may indicate an impairment in the neural processes that promote one percept while suppressing the other, which could reflect disrupted E/I balance. Prior research has shown that switch rates during binocular rivalry are slower in PwPP relative to healthy controls (Fox, 1965; Miller et al., 2003; Nagamine et al., 2009; Ngo et al., 2011; Xiao et al., 2018; Ye et al., 2019). Additionally, studies using MR spectroscopy in PwPP have found abnormalities in neural metabolite concentrations in visual cortex, including glutamate and GABA (Marsman et al., 2014; Natsubori et al., 2014; Sydnor & Roalf, 2020; Wenneberg et al., 2020; Yoon et al., 2010). However, the specific role of different neurotransmitters during bi-stable perception in psychosis is currently unknown.

Genetics is another factor known to play a significant role in the etiology of psychotic disorders, including schizophrenia and bipolar disorder (Cardno et al., 1999; Cardno & Owen, 2014). One way to study the role of genetic liability for psychosis in perceptual and cognitive dysfunction is to study task performance among unaffected biological relatives of PwPP (Keri et al., 2001; M. P. Schallmo et al., 2015; Silverstein et al., 2006; Sponheim et al., 2013). First-degree biological relatives (i.e., parents, siblings, and children) share on average 50% of their genes with PwPP. If abnormal perceptual or cognitive performance is observed among such relatives, this may provide evidence for an endophenotype (Calkins et al., 2008; Gottesman & Gould, 2003; Iacono et al., 2017), which is an anomaly associated with genetic liability for psychosis, rather than with a diagnosed psychotic disorder per se.

In the current study, we tested the hypothesis that PwPP have impaired visual integration of local motion cues into a global percept. Specifically, we examined bi-stable perception in PwPP, their close biological relatives, and healthy controls using a SFM rotating cylinder task. We also measured metabolite concentrations in visual cortex using MR spectroscopy at 7 tesla, including glutamate, GABA, and glutamine levels, to examine the potential role of these metabolites during SFM perception in PwPP. Based on previous studies of binocular rivalry (Fox, 1965; Xiao et al., 2018; Ye et al., 2019), we expected to find slower switch rates during SFM perception in PwPP. Contrary to our prediction, we in fact observed *faster* switch rates during SFM perception in both PwPP and their biological relatives, compared to healthy controls. Our findings suggest an impairment in bi-stable visual motion and form perception that is linked to genetic liability for psychosis.

## 2. Methods

This study was conducted as part of a larger series of experiments in the Psychosis Human Connectome Project (pHCP). The essential details of our experimental approach are described below. For a full description of all pHCP study methods, please see (Demro et al., 2021; Schallmo et al., 2023).

### 2.1 Participants

Participants were recruited as part of the pHCP study at the University of Minnesota (Table 1). Data were collected as part of a series of experiments focused on visual perception, which included fMRI and MR spectroscopy at 7 tesla (Schallmo et al., 2023).

**Table 1.**
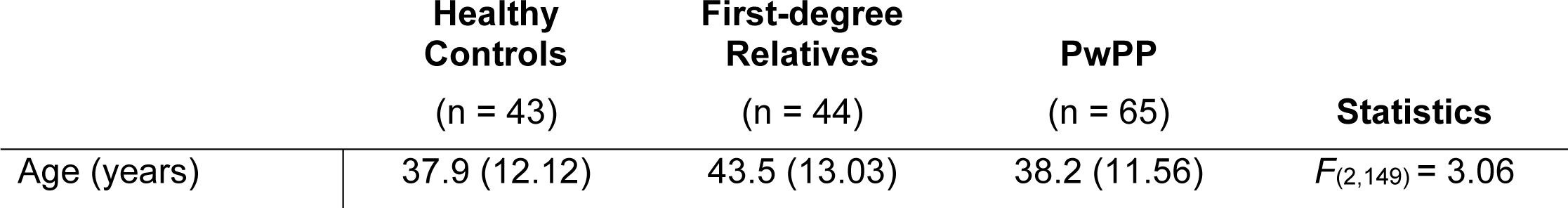
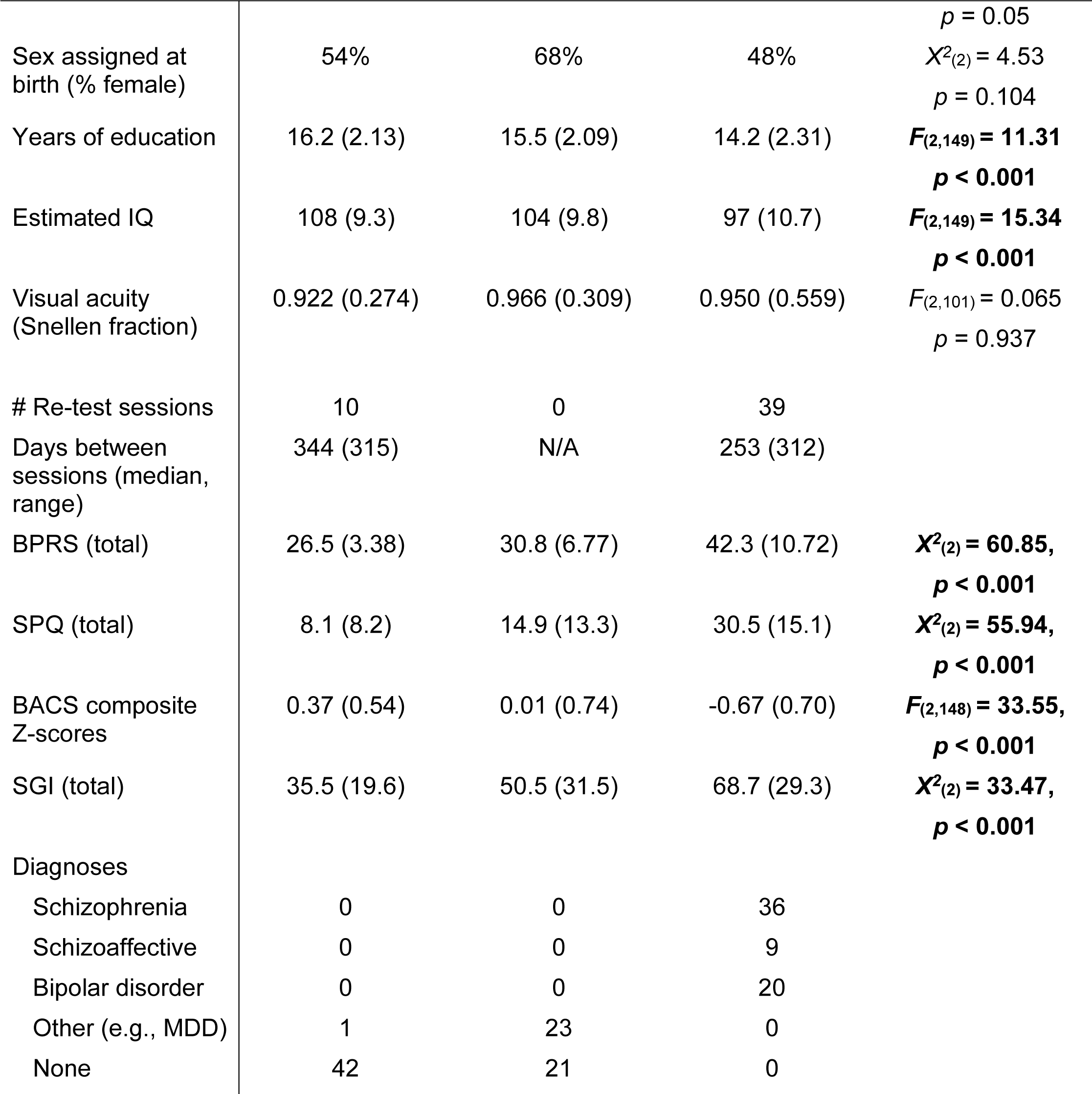
Subject group demographics, clinical symptom levels, and cognitive measures. Data are presented as mean (standard deviation), unless otherwise specified. Estimated IQ was measured using the Wechsler Adult Intelligence Scale (WAIS-IV; Wechsler, 2008). Visual acuity was measured using a Snellen visual acuity test (Snellen, 1862). The Snellen fraction is reported (e.g., 1 indicates 20/20 vision). The clinical tests administered were: BPRS (Brief Psychiatric Rating Scale - Total Score; Overall & Gorham, 1962), SPQ (Schizotypal Personality Questionnaire; Raine, 1991), and BACS (Brief Assessment of Cognition in Schizophrenia – Z-score; Keefe et al., 2004). The statistics column shows the test statistics and *p*-values calculated across the three groups for each measure. Highlighted entries indicate significant group differences at *p* < 0.05. For any measure in which normality and / or homogeneity of variance were not observed, non-parametric Kruskal-Wallis tests (*X^2^*) were used in place of ANOVAs (*F*).

A total of 152 participants participated in the SFM task. Participants were divided into three groups: people with a history of psychotic psychopathology (PwPP hereafter), their first-degree biological relatives (relatives hereafter), and healthy controls (see Table 1 for detailed demographics). Of the 152 participants initially tested, 49 returned for a second (re-test) session (Table 1). The median number of days participants returned after their initial session was 133.5 (range: 36-1173 days; see Supplemental Figure 1).

All participants were between 18-65 years of age, spoke English as their primary language, provided written and informed consent, had not been diagnosed with any learning disability, did not have an IQ of less than 70, nor any current or past central nervous system disease. Participants in the psychosis group had a history of a disorder with psychotic psychopathology (i.e., schizophrenia, schizoaffective, bipolar), whereas relatives were a biological parent, sibling, or offspring of the individuals with psychotic psychopathology. All participants had Snellen visual acuity (with correction, if used) of 20/40 or better. Individuals with poorer than 20/40 acuity were excluded. A total of 3 participants were excluded based on the criteria above (0 controls, 2 relatives, 1 PwPP) and are not included in Table 1. As part of the pHCP participants participated in two 3 T fMRI scanning sessions and one or two 7 T scanning sessions. Details of the clinical assessments and the 3 T scanning protocol are included in our recent publication (Demro et al., 2021). SFM task data were collected outside of the scanner, in a separate psychophysics room, prior to 7 T scanning on the same day. All participants provided written informed consent prior to participating and were compensated $20 per hour. All procedures were in compliance with the Declaration of Helsinki and were approved by the University of Minnesota IRB.

### 2.2 Apparatus

Psychophysical data were collected at the Center for Magnetic Resonance Research at the University of Minnesota. Data were collected on an Apple Mac Pro using an Eizo FlexScan SX2462W monitor with a 60 Hz refresh rate (mean luminance = 61.2 cd/m^2^). A Bits# stimulus processor (Cambridge Research Systems) was connected but not used for this particular experiment. Luminance values were linearized using a PR655 spectrophotometer (Photo Research). Head position was stabilized using an adjustable chin rest positioned at a viewing distance of 70 cm. Stimuli were generated and responses were collected using PsychoPy (Peirce et al., 2019).

### 2.3 Stimuli

The visual stimuli used were standard structure-from-motion rotating cylinders (Treue et al., 1991; Ullman, 1979). The rotating cylinder is a classic illusion in which small moving elements (here, black and white squares) move back and forth across a rectangular area in order to induce the perception of a cylinder rotating in 3-dimensional space. The rotating cylinder stimuli used here were composed of 400 small black and white squares (each 0.25° of visual angle; 200 black and 200 white) that alternated between moving from the left to right and right to left across the width of a rectangular area and each positioned pseudo-randomly along the height of the rectangular region (height = 10°, width = 7°). The squares sped up as they approached the center of the rectangle and slowed down as they approached each edge. This was done to simulate the perceived speed of the squares as if they were positioned on a transparent cylinder rotating in the depth plane (simulated rotation speed of 90°/s).

Two versions of the stimuli with subtle but important differences were used for the two different task conditions, referred to as the real switch and bi-stable tasks. In both tasks, the same black and white squares moved back and forth across the rectangular area with speed, size, and position being constant across the two tasks. In the bi-stable task, a small red fixation point (0.6° diameter) was positioned in front of all squares in the center of the rectangular area (Figure 1A). When dots collided in this task, they occluded one another randomly in order to remove this potential depth cue. In the absence of inherent depth cues, the direction of motion is ambiguous, and perception spontaneously alternates between the front surface rotating to the left and to the right.

**Figure 1.**
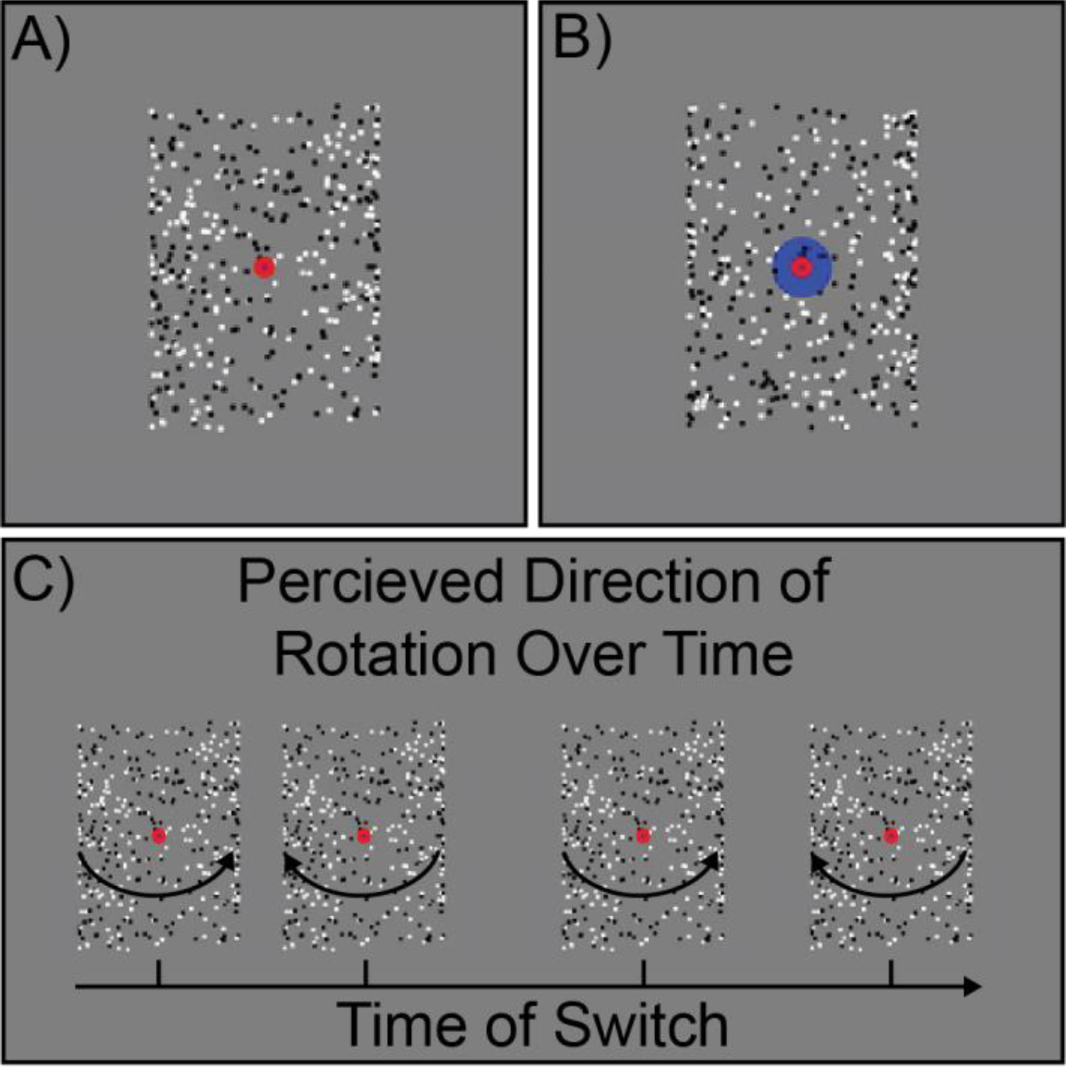
Stimulus and task design. Static examples of (A) the bi-stable and (B) real switch task stimuli. C) Hypothetical time course of rotation switches during the bi-stable task. Figure reproduced from (Schallmo et al., 2023).

In the real switch task, we simulated physical switches of the rotation direction in depth by changing which set of dots (i.e., those rotating left to right or right to left, surface #1 and surface #2 of the cylinder) were presented in the front or back. To do so, we used the same red fixation dot overlaid on a larger blue circle (1.8° diameter) and alternated which set of dots passed in front of or behind the larger fixation circle (Figure 1B). This provided an explicit depth cue, thus biasing one percept (e.g., front rotating to the left) to become dominant. Switches occurred every 9-13 seconds with a total of 11 real switches per block (order and timing was pseudorandomized, but fixed and identical for all subjects). The real switch task was added to the experiment in October 2017, thus data in this condition were not collected from a number of the early data sets (10 controls, 0 relatives, 2 PwPP). Of these, 1 control and 2 PwPP were excluded based on other exclusion criteria listed in the Participants section above, leaving 9 control data sets with missing real switch task data. Four of these controls returned for a re-test session during which real switch task data were collected (none of which were excluded for poor real switch task data quality, see below). We chose to retain the bi-stable task data from participants who did not have real switch task data, which gave us different numbers of control participants for the two tasks.

### 2.4 Experimental procedure and task design

In both the real switch and bi-stable tasks, participants were asked to fixate on the small red central circle and use their peripheral vision to determine the direction of rotation of the front surface of the cylinder, either left or right. Participants were then instructed to respond using the left or right arrow keys to indicate which direction of rotation they perceived. Importantly, they were told to respond immediately at the beginning of the stimulus presentation to their initial percept, and then again each time their perception changed. Each participant first completed a short 30 s practice version of the real switch task to familiarize themselves with the task. They then ran one block of the real switch task and 5 blocks of the bi-stable task. Each block was 2 min long; the rotating cylinder was presented for the entirety of the block.

### 2.5 Behavioral task analysis

#### 2.5.1 Real Switch Task

Data from the real switch task were used to exclude subjects who had trouble detecting real stimulus direction switches from further analyses (i.e., of their bi-stable task data). We defined correct responses in the real switch task as those that matched the stimulus rotation direction and occurred within 4 s after a direction change. This was based on our examination of the average reaction time for correct responses (Supplemental Figure 2), the average percept durations towards and away from the physical rotation direction (Supplemental Figure 3), the distribution of reaction times during the real switch task (Supplemental Figure 4), and the total number of reported switches (Supplemental Figure 5). We defined poor real switch task performance as having 6 or fewer correct responses (≤ 63.6% accuracy) made within 4 s of a stimulus direction change (Supplemental Figure 4B). We chose these thresholds after visually inspecting the data and determining that the large majority of participants and responses fell above these thresholds. Of the 147 total data sets with real switch data, including re-test sessions, we excluded a total of 30 data sets who did not meet the above *post-hoc* criteria (6 controls, 7 relatives, 17 PwPP).

#### 2.5.2 Bi-stable Task

In order to assess the stability of ambiguous motion direction percepts in this task, we compared the distribution of durations for all reported percepts across the three groups using two sample Kolmogorov-Smirnov tests. This allows us to examine how the durations of reported percepts may have differed between groups.

Next, we calculated the switch rate (switches reported per second) and average percept duration (average length of percept dominance) during each task for each subject. To quantify switch rates, we measured the total number of switches in each block and divided by the total time (2 min per block). Because the switch rate data were highly skewed (based upon visual inspection), we then normalized the data by performing a log_10_ transformation. As it is possible to not respond during a 2 min block in the bi-stable task, some participants can have switch rates of 0 Hz, which when normalized using log_10_, become values of negative infinity. Therefore, we replaced values of 0 switches per block with 0.5 switches / 120 s when normalizing. This occurred for a total of 2 participants (1 PwPP, 1 control). Although we measured both switch rate and percept duration, both of these measures displayed the same trends, and were highly correlated with one another.

Therefore, to avoid repetition, we present results of the switch rate analysis in the main text, while percept duration data are presented in the Supplemental Information. We note that although the switch rate and percept duration data were very similar, they were not identical. This is because the percept duration is calculated only after the participant’s initial response in each block, which occurs a short time after the stimulus onset, whereas the switch rate was defined based on the full two minute duration of the block. As the data in both the real switch and bi-stable tasks did not have equal variance across groups and were skewed (even after log_10_ transformation), we performed a Kruskal-Wallis one-way non-parametric ANOVAs to assess group differences in switch rates during the real switch task.

We also measured the stability of bi-stable perception dynamics over time by examining longitudinal variability in a subset of our participants. To do so, we compared switch rates in the bi-stable task measured across two different experimental sessions (months apart) within the same individuals (49 individuals with re-test data; 10 controls, 0 relative, 39 PwPP). Of these individuals with re-test data, 9 participants (1 control and 8 PwPP) had 1 or more of their data sets excluded based on poor real switch task performance. This left a total of 40 individuals (9 controls and 31 PwPP) with usable re-test data. Information about the amount of time between testing sessions and number of participants who completed a second test session is provided in Table 1. We calculated the intraclass correlation coefficient (ICC(3,k)) between switch rates for the first and second sessions (Shrout & Fleiss, 1979).

Finally, we sought to identify any differences between clinical diagnoses within our PwPP group. Specifically, we compared participants with schizophrenia (n = 25) and bipolar disorder (n = 16) to healthy controls (n = 37). People with schizoaffective disorder were not included in this analysis, due to a smaller sample size (n = 7).

### 2.6 Association between clinical symptom level and switch rate

We used a set of measures to assess psychiatric symptoms and cognitive functioning, which included the Schizotypal Personality Questionnaire (SPQ; Raine, 1991), The Brief Assessment of Cognition in Schizophrenia; (BACS; Keefe et al., 2004), The Brief Psychiatric Rating Scale (BPRS; Overall & Gorham, 1962), and The Sensory Gating Inventory (SGI; Bailey et al., 2021; Hetrick et al., 2012). Out of a variety of clinical assessments administered as a part of the pHCP (Demro et al., 2021), we chose these as they provide measures of overall psychiatric symptom levels, schizotypy, and cognitive functions, and previous work has found relationships between these factors and visual dysfunction in people with psychotic disorders (Schmack et al., 2015; Weilnhammer et al., 2020; Xiao et al., 2018). These measures were collected for participants in all three groups (unlike other measures that were collected only from PwPP). In addition to the four primary measures mentioned here, we also chose to include the cognitive-perceptual subscale from the SPQ (SPQ-CP) and a disorganization factor from the BPRS (BPRS-D; Wilson & Sponheim, 2014), as they are particularly relevant to our analyses and hypotheses.

To investigate relationships between symptom severity and percept switch rate, we first correlated individual symptom measures with average switch rates across all three groups. We used Spearman rank correlations to avoid assuming linear relationships. Data from re-test sessions were excluded from these correlations, as Spearman correlations assume independent data points.

### 2.7 Quantification and analysis of neurotransmitter concentrations from MR spectroscopy

Neurochemical concentrations in the mid-occipital lobe were collected as part of a 7 T magnetic resonance spectroscopy (MRS) scan on the same day as behavioral SFM data. For full scanning details, see (Schallmo et al., 2023). Data were acquired on a Siemens MAGNETOM 7 T scanner with a custom surface radio frequency head coil using a STEAM sequence (Marjańska et al., 2017) with the following parameters: TR = 5000 ms, TE = 8 ms, volume size = 30 mm (left-right) x 18 mm (anterior-posterior) x 18 mm (inferior-superior), 3D outer volume suppression interleaved with VAPOR water suppression (Tkáč et al., 2001), 2048 complex data points with a 6000 Hz spectral bandwidth, chemical shift displacement error = 4% per ppm. B_0_ shimming was performed using FAST(EST) MAP to ensure a linewidth of water within the occipital voxel ≤ 15 Hz (Gruetter, 1992).

We processed our MRS data using the *matspec* toolbox (<u>github.com/romainVala/matspec</u>) in MATLAB, including frequency and phase correction. Concentrations for 18 different metabolites including glutamate, glutamine, and GABA were quantified in each scanning session using LCModel. We scaled metabolite concentrations relative to an unsuppressed water signal reference, after correcting for differences in gray matter, white matter, and CSF fractions within each subject’s MRS voxel, the proportion of water in these different tissue types, and the different T_1_ and T_2_ relaxation times of the different tissue types. Tissue fractions within the voxel were quantified in each subject using individual gray matter and white matter surface models from FreeSurfer (Fischl, 2012). MRS data sets were excluded based on the following data quality criteria: H_2_O line width > 15, LCModel spectrum line width > 5 Hz or LCModel SNR < 40. Out of a total of 193 MRS datasets (54 controls, 44 relatives, and 95 PwPP), 10 sets (1 control, 4 relatives, 5 PwPP) were excluded in this way, leaving 183 total MRS datasets. In addition to subjects whose SFM data we excluded for having poor real switch task performance, and excluding re-test sessions, this left a total of 114 participants with usable SFM and MRS data (37 controls, 33 relatives, and 44 PwPP).

In order to probe the possible role of excitatory and inhibitory markers during bi-stable perception in PwPP, we examined relationships between metabolite concentrations from MRS and our bi-stable SFM behavioral measures. Specifically, we used Spearman rank correlation to test for correlations between metabolite levels from MRS (i.e., GABA, glutamate, and glutamine) and average switch rates across participants from all three groups. As in our other correlational analyses, data from retest sessions were excluded, as Spearman correlations assume independence across data points.

## 3. Results

### 3.1 Individuals with psychotic psychopathology reliably detect real switches in rotation direction

To ensure that all participants understood the task and were able to detect real changes in rotation direction, we used a real switch task in which physical depth cues were present. This allowed us to measure detection of real rotation direction switches.

We examined the distributions of the number of responses made in the real switch task in each group (Supplemental Figure 5; panel A shows all responses, B shows responses with reaction time < 4 s), to ensure participants were responding to the majority of physical switches. The large majority of participants responded to ≥ 63.4% of the physical switches or 7/11 total switches with a reaction time < 4 s (83.3% controls, 81.4% relatives, and 70.7% PwPP). We excluded a total of 30 individual participants who did not meet the above *post-hoc* criteria (6 controls, 7 relatives, 17 PwPP), leaving a total of 122 participants with bi-stable task data. To determine whether exclusion rates differed across groups, we performed a contingency table analysis. The rate of excluded participants across groups was not significantly different from what would be expected by chance (*X^2^*_(2)_ = 1.55, *p* = 0.46).

As can be seen in Figure 2, PwPP did not differ significantly in real switch task accuracy (*X*^2^_(2)_ = 5.32, *p* = 0.069), as compared to healthy controls or first-degree biological relatives of PwPP. However, there was a significant difference in reaction time in the real switch task (*X*^2^_(2)_ = 9.33, *p* = 0.009; Supplemental Figure 2), with PwPP tending to respond most slowly. Importantly, our analyses of the real switch task data demonstrate that participants in this study, including PwPP, can accurately detect real switches in direction for the rotating cylinder. This suggests that participants in each group generally understood the task instructions, could integrate the local stimulus elements into a global rotational motion percept, and could correctly respond to physical changes in depth cues (i.e., occlusion) to disambiguate the cylinder’s rotation direction.

**Figure 2.**
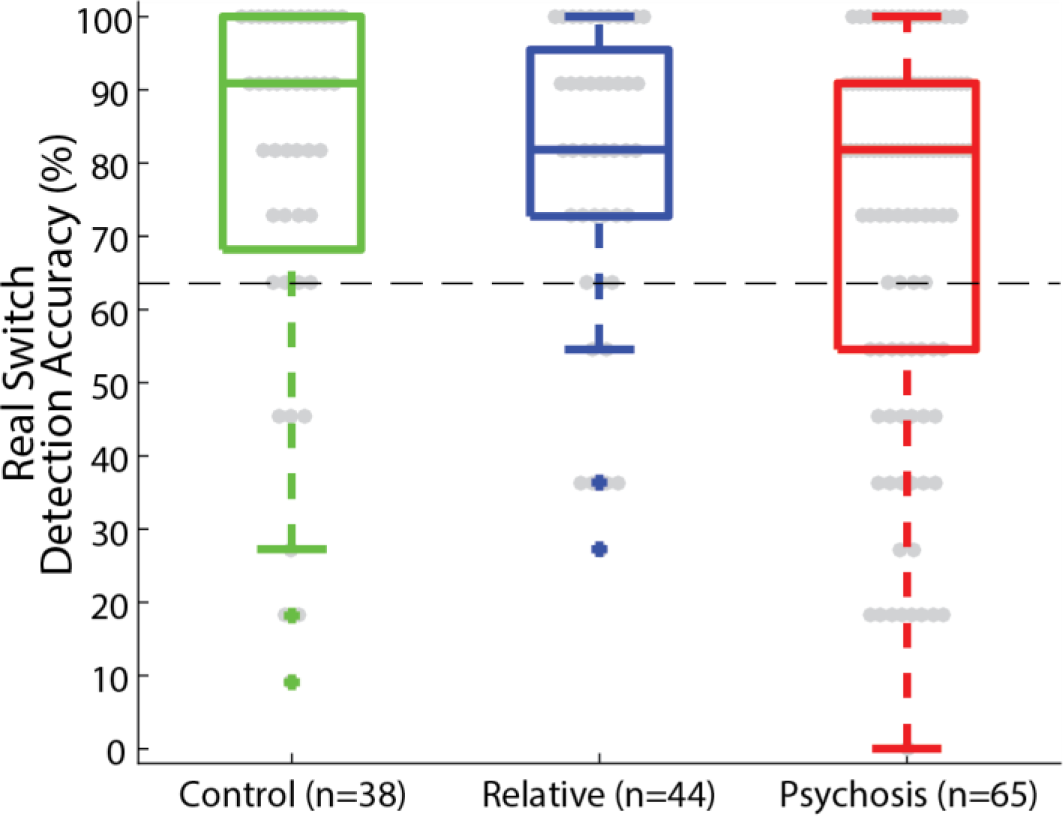
Real switch task results. Box plots depict the median accuracy (middle line) as well as the 25^th^ and 75^th^ quartiles (box), 1.5 x the interquartile range (whiskers), and outliers (plus) for the real switch task for each group (controls: green, relatives: blue, psychosis: red). Data plotted represent detection accuracy for all correct responses (those that matched the real rotation direction, and that were made within 4 s of the direction change). The dashed horizontal line represents the exclusion cutoff value of ≥ 63.6% accuracy.

### 3.2 Faster switch rates in individuals with psychotic psychopathology

We hypothesized that integration of local information into a global percept is impaired for individuals with psychotic psychopathology, which results in abnormal bi-stable switch rates. To examine this, we first looked at histograms of the distributions of reported percept durations in each group, in order to test whether the three groups reported similar switch dynamics. As shown in Figure 3A & B, although the distributions of percept durations from the three groups follow similar trends, they are significantly different (Kolmogorov-Smirnov (K-S) test real switch task – controls vs. relatives: *D*_(32,37)_ = 0.146, *p* < 0.001; controls vs. PwPP: *D*_(32,48)_ = 0.281, *p* < 0.001; relatives vs. PwPP: *D*_(37,48)_ = 0.147, p < 0.001; K-S test bi-stable task - controls vs. relatives: *D*_(37,37)_ = 0.082, *p* < 0.001; controls vs. PwPP: *D*_(37,48)_ = 0.13, *p* < 0.001; relatives vs. PwPP: *D*_(37,48)_ = 0.075, *p* < 0.001). For the real switch task (Figure 3A), this indicates that participants in all 3 groups could detect the real switches occurring at intervals of 9, 11, and 13 s, but that relatives, and especially PwPP, tended to report shorter percept durations. For the bi-stable task (Figure 3B), distributions were also shifted toward shorter percept durations for PwPP versus controls, with relatives showing an intermediate pattern.

**Figure 3.**
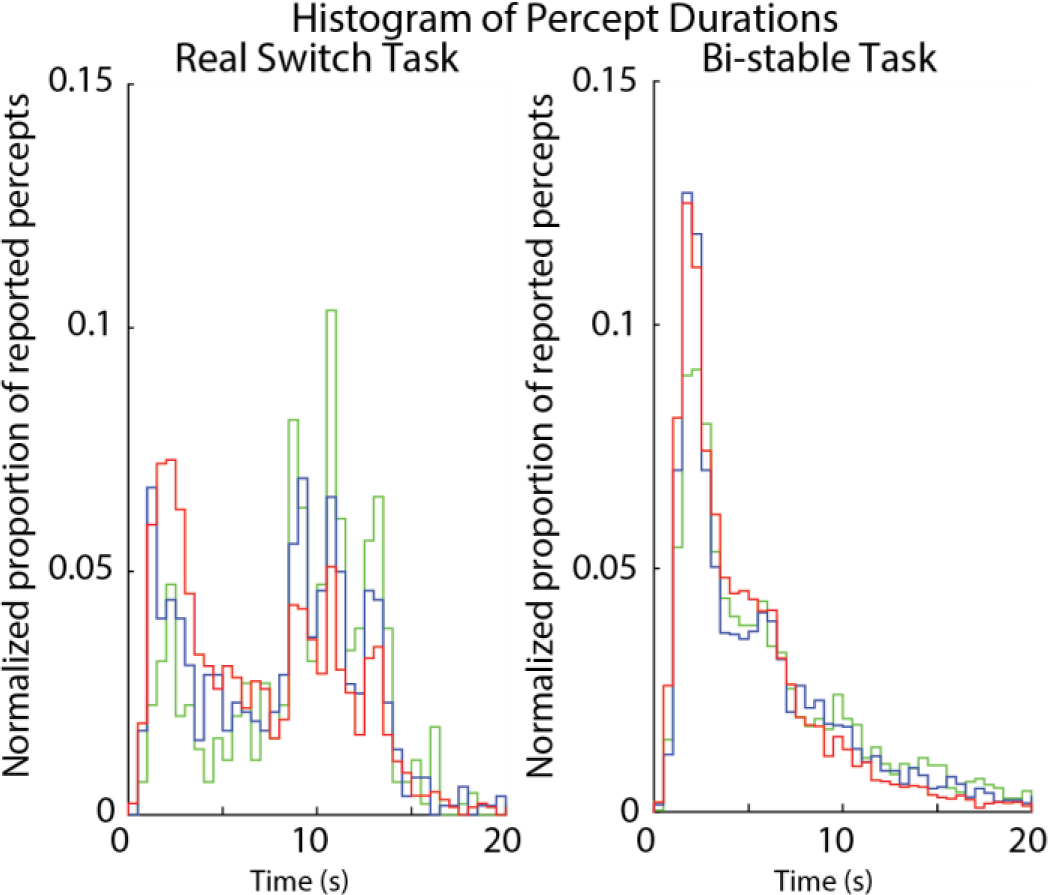
Histograms of percept durations for (A) the real switch and (B) bi-stable tasks. Shown are normalized distributions of percept durations, defined as times between reported switches, across the three groups (controls, n = 32 & 37 – green, relatives, n = 37 & 37 – blue, psychosis, n = 48 & 48 – red). Distributions were normalized by dividing by the total number of responses across all participants within each group.

Next, we examined switch rates for both the real switch and bi-stable tasks (Figure 4). The real switch task data allowed us to assess how well participants could detect real switches in rotation direction based on explicit depth cues, whereas the bi-stable task probed spontaneous percept switches in the absence of such cues.

**Figure 4.**
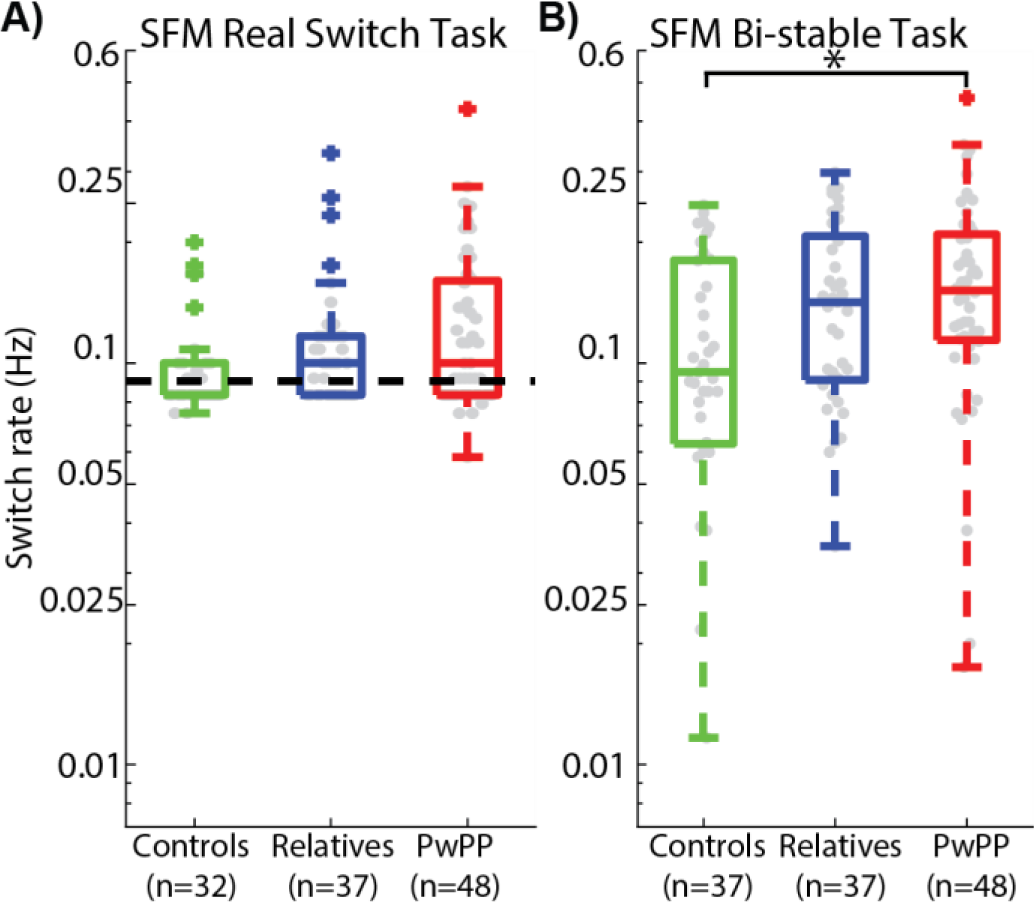
Switch rates for (A) the real switch task and (B) bi-stable task. Box plots show median (middle line), 25%-75% quartiles (box), 1.5 x the interquartile range (whiskers) and outliers (plus) for the three groups. Gray dots show average switch rates from individual participants. Dashed line in A shows average switch rate for physical stimulus changes in the real switch task. Bi-stable switch rates (B) were significantly higher among PwPP vs. controls (*).

During the real switch task PwPP, relatives, and controls tended to report perceptual switches at similar rates (Figure 4A). As described in the Methods section, the real switch task had physical switches overtly implemented that occurred at an average rate of 0.09 Hz (i.e., every 9-13 s). There was no significant group difference in switch rates in the real switch task (*X^2^*_(2)_ = 5.36, *p* = 0.069), and median values across participants were close to the true physical switch rate, indicating that participants in each group generally could detect real switches in rotation direction in this task. This suggests that all participant groups were able to integrate local motion signals into an unambiguous global percept of rotational motion given some explicit depth cues.

During the bi-stable task, we observed a significant difference in switch rates across groups (*X^2^*_(2)_ = 6.49, *p* = 0.039), with PwPP and their relatives having faster switch rates overall versus controls (Figure 4B). Post-hoc comparisons showed that switch rates among PwPP were significantly higher than in controls (*X*^2^_(1)_ = 6.93, *p* = 0.0085). This suggests that the ability to integrate ambiguous motion stimuli into a global percept in the absence of contextual depth information may be disrupted among PwPP.

Switch rates for the relative group also differed significantly from controls but not from PwPP (controls vs. relatives: *X*^2^_(1)_ = 3.98, *p* = 0.046; PwPP vs. relatives: *X*^2^_(1)_ = 0.516 *p* = 0.473). This suggests that genetic risk for psychotic disorders, as expressed by unaffected first-degree relatives, may be associated with an attenuated disruption in bi-stable motion perception during our SFM task.

Additionally, we found a main effect of time (i.e., block number: *F*_(119,2)_ = 27.15, *p* < 0.001) in the bi-stable task, indicating a slight overall decrease in percept switch rates across the five 2-min task blocks. As can be seen in Supplemental Figure 6, the pattern of differences in switch rates between the three groups remained consistent across blocks. Changes in switch rates over time did not differ significantly between groups (*F*_(119,2)_ = 0.1.046, *p* = 0.354). The effect of slower switch rates during later task blocks may reflect an effect of practice, learning, or adaptation during bi-stable task performance.

We measured both switch rates and percept durations in the bi-stable task; both of these measures displayed the same trends across groups (group difference in percept duration, *X^2^*_(2)_ = 8.67, *p* = 0.013; controls vs. PwPP: *X*^2^_(1)_ = 7.45, *p* = 0.0063) and were highly correlated with one another. Likewise for the real switch task, we saw no significant group differences in percept duration (*X^2^*_(2)_ = 2.85, *p* = 0.24). We show the average percept duration results in Supplemental Figure 7.

In order to assess whether variability in bi-stable perception differed across groups, we calculated the coefficient of variance (i.e., standard deviation of percept duration divided by the mean) for both task types within each participant. We found no significant difference in percept variability between groups (real switch: *X*^2^_(2)_ = 5.351, *p* = 0.067; bi-stable: *X*^2^_(2)_ = 4.06, *p* = 0.131; see Supplemental Figure 8), suggesting that the variability of percept durations was generally comparable across groups.

Lastly, we analyzed differences in bi-stable perception across different clinical diagnoses. We compared switch rates among people with schizophrenia, people with bipolar disorder, and healthy controls (Supplemental Figure 9). Unlike in the full group comparisons, we did see significant differences between these diagnostic groups for the real switch task (*X^2^*_(2)_ = 6.85, *p* = 0.033). Post-hoc comparisons between the individual groups showed a significant difference between the control and bi-polar groups (controls vs. bipolar disorder: *X*^2^_(1)_ = 4.34, *p* = 0.037). This suggests that patients with bipolar disorder may have had particular difficulty in detecting real switches. However, we did not see any differences between the schizophrenia group and healthy controls (controls vs. schizophrenia: *X*^2^_(1)_ = 1.45, *p* = 0.228) or between the two psychosis groups (schizophrenia vs. bipolar disorder: *X*^2^_(1)_ = 0.818, *p* = 0.366). For the bi-stable task we once again saw a significant effect of group, where people with schizophrenia and bipolar disorder switched at a faster rate than controls (*X^2^*_(2)_ = 11.28, *p* = 0.004). Post-hoc comparisons showed no differences between people with bipolar disorder and healthy controls (controls vs. bipolar disorder: *X*^2^_(1)_ = 2.52, *p* = 0.112) or between the two psychosis groups (schizophrenia vs. bipolar disorder: *X*^2^_(1)_ = 0.086, *p* = 0.769). However, there was a significant difference between people with schizophrenia and healthy controls (controls vs. schizophrenia: *X*^2^_(1)_ = 5.304, *p* = 0.021). As can be seen in Supplemental Figure 9, these results suggest that differences in switch rates between the diagnostic sub-groups and controls may be smaller / less reliable than for the psychosis groups as a whole. Alternatively, we may have had insufficient statistical power to detect diagnostic group differences, given the smaller number of participants in each sub-group.

### 3.3 Switch rates remain consistent over months

Next, to examine the stability of bi-stable perception dynamics over a long period of time, we re-tested a subset of participants in a second experimental session. Table 1 and Supplemental Figure 1 show the median and range of the time between the first and second sessions in each group (median = 133.5 days). Across participants, the inter-class correlation for switch rates during the first and second session was high (ICC(3,k)=0.88; Figure 5), indicating that switch rates remained rather consistent within participants over a period of several months. Additionally, this provides evidence that abnormalities in bi-stable perception among the psychosis group are relatively stable over a long time period.

**Figure 5.**
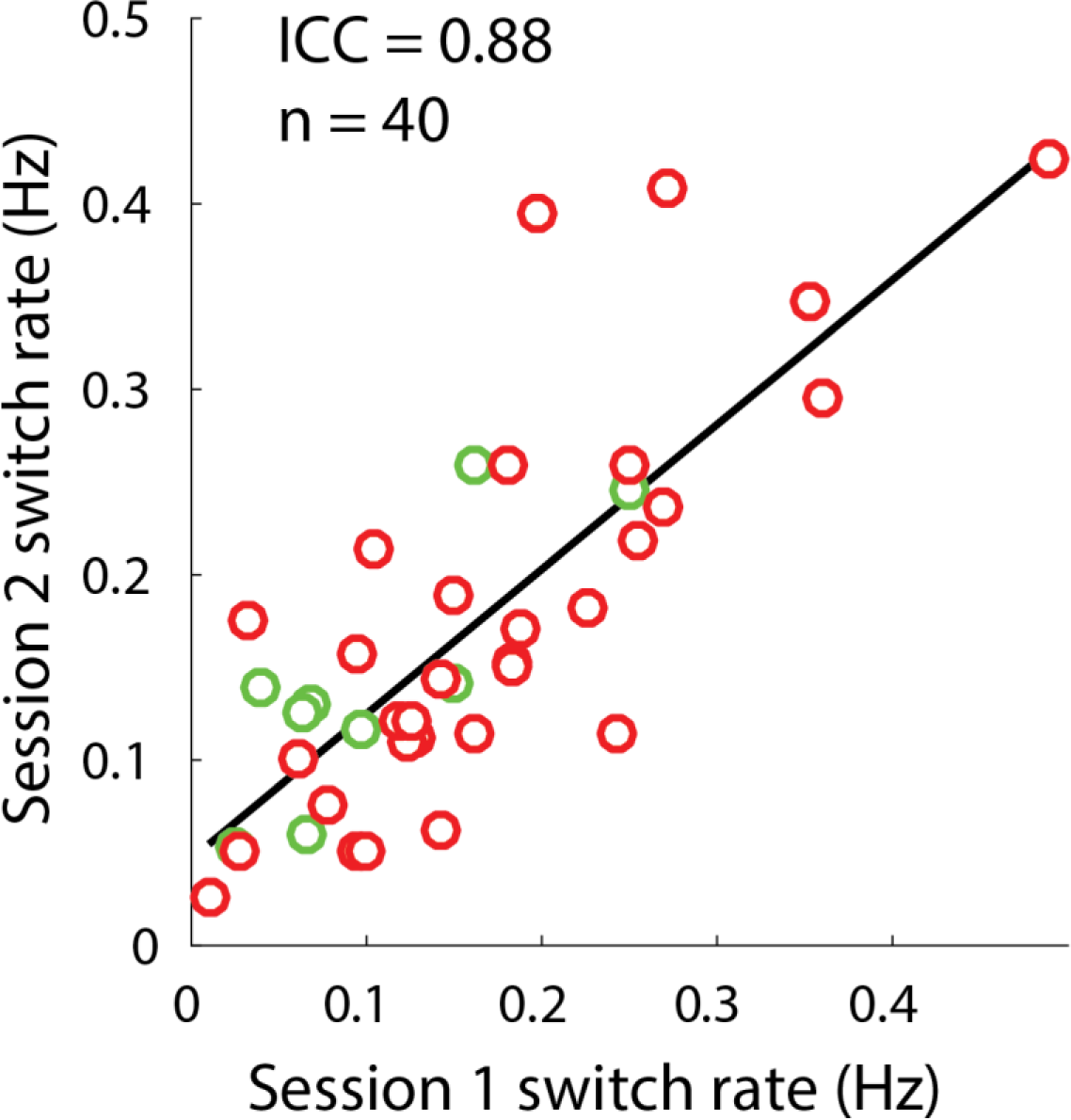
Longitudinal variability of bi-stable switch rates. Plotted are switch rates (Hz) for session 1 (x-axis) and session 2 (y-axis) for all subjects who participated in two sessions. Red dots show data from PwPP (n = 31), green dots show data from controls (n = 9).

### 3.4 Association between bi-stable perception and clinical symptoms

We next examined the relationships between measures of clinical psychopathology and bi-stable perception. To do so, we quantified Spearman correlations between clinical scores and bi-stable switch rates (Supplemental Figure 10). We found no significant relationships between switch rates and measures of cognitive functioning (BACS; *r*_(119)_ = -0.139, uncorrected *p* = 0.129) or sensory gating (SGI; *r*_(120)_ = 0.161, uncorrected *p* = 0.077). However, we did find a correlation between faster switch rates and higher overall psychiatric symptom scores (BPRS; *r*_(103)_ = 0.207, uncorrected *p* = 0.034) as well as schizotypy (SPQ; *r*_(120)_ = 0.188, uncorrected *p* = 0.038). We also examined the cognitive-perceptual subscale of the SPQ and a disorganization factor from the BPRS (Wilson & Sponheim, 2014) as these measures are relevant to cognitive / perceptual dysfunction. We found positive relationships with switch rate for both measures (BPRS-D; *r*_(103)_ = 0.264, uncorrected *p* = 0.006, Bonferroni corrected *p* = 0.036; SPQ-CP; *r*_(120)_ = 0.209, uncorrected *p* = 0.021). This indicates that across all groups, individuals who had higher levels of psychiatric symptoms, especially disorganization, also showed greater perceptual abnormalities in terms of faster bi-stable switch rates. However, of these, only the BPRS-D correlation survived correction for multiple comparisons. Thus, these findings may be considered preliminary and in need of confirmation.

### 3.5 Neurotransmitter concentrations and perceptual bi-stability

We quantified concentrations in the medial occipital lobe using 7 T ^1^H-MRS for a number of metabolites, including GABA, glutamate, and glutamine. We chose to focus our analyses on these 3 metabolites as they are putative excitatory and inhibitory markers and have, in the case of GABA, previously been shown to correlate with SFM switch rates among healthy adults (van Loon et al., 2013). First, we compared concentrations across our three groups for each of these three metabolites (Supplemental Figure 11). We did not observe any significant differences across the three groups for these three metabolites (GABA: *X*^2^_(2)_ = 0.194, *p* = 0.91; glutamate: *X*^2^_(2)_ = 0.80, *p* = 0.67; glutamine: *X*^2^_(2)_ = 1.07, *p* = 0.58). We then quantified correlations between metabolite concentrations and switch rates from the bi-stable task across all participants, excluding repeated scan data (Supplemental Figure 12). We found no significant relationship between switch rates and occipital concentrations of glutamate (*r*_(112)_ = -0.058, *p* = 0.54) or glutamine (Spearman’s: *r*_(112)_ = 0.06, *p* = 0.53). We did see a non-significant trend between higher occipital GABA levels and slower SFM switch rates (*r*_(106)_ = -0.16, *p* = 0.095), which is in the same direction as a relationship observed in healthy adults that has been reported previously (van Loon et al., 2013). However, as our result does not reach statistical significance, this finding should be interpreted with caution. Correlations between GABA levels and switch rates within each group were not significant for both controls (*r*_(35)_ = -0.091, uncorrected *p* = 0.59) or PwPP (*r*_(42)_ = -0.091, uncorrected *p* = 0.558). There was a correlation between higher GABA levels and slower switch rates within the relatives group, but it did not survive correction for multiple comparisons (*r*_(31)_ = -0.367, uncorrected *p* = 0.036, Bonferroni corrected *p* = 0.11). Thus, we did not find strong evidence from MRS to support a relationship between excitatory or inhibitory markers in visual cortex and switch rates from our bi-stable SFM task.

## 4. Discussion

We examined visual motion and form integration across the psychosis spectrum using a bi-stable SFM perception task among PwPP, their first-degree relatives, and healthy controls. In our bi-stable task, we found significantly faster switch rates among PwPP compared to healthy controls, with relatives showing an intermediate pattern of results. Faster switch rates in these groups suggest that SFM percepts were less robust or stable during the task. The level of instability tended to be consistent across experimental sessions held on different days several months apart. There were no significant group differences in switch rates during our real switch task, indicating a similar ability to detect actual physical changes in rotation direction. Across participants, faster switch rates correlated with significantly higher psychiatric symptom levels, implying that this perceptual abnormality co-varied with the degree of mental health symptoms. Together, our findings indicate that current psychotic illness is associated with abnormal perception of the bi-stable rotating cylinder illusion, while unexpressed genetic liability for psychosis may also be associated with this perceptual abnormality.

Prior research has shown that processing of the rotating cylinder requires processing of both object shape (Freeman et al., 2012; Murray et al., 2002) and complex rotational motion (Braddick et al., 2000; Rees et al., 2000; Smith et al., 2006), and involves brain regions implicated in both processes, such as LOC and hMT+. Therefore, differences in switch rate among PwPP may be caused by impaired integration of local motion cues into a larger rotating object percept. Consistent with this notion, previous studies have found that visual functions involving integration (including center-surround suppression, contour integration, border detection, object and shape representation, figure-ground segmentation, and structure-from-motion) are also impaired in PwPP (Carter et al., 2017; Chen et al., 2003, 2005, 2008; Dakin et al., 2005; Keane et al., 2012; Pokorny et al., 2020; Robol et al., 2013; M. P. Schallmo et al., 2013, 2015; Silverstein et al., 2006; Tadin et al., 2006).

Our results suggest that the faster switch rates we observed in our SFM task among PwPP may not be attributed solely to impaired local motion detection, but rather involve impairments in the ability to combine those local motion signals into one of two stable competing percepts of a rotating cylinder. Performance in our real switch task depends on the ability to integrate local motion cues to detect rotation direction, and the explicit depth information facilitates the perception of an unambiguous global motion percept. Although we observed differences across groups in the distributions of percept durations during our real switch task (i.e., greater numbers of shorter percepts in PwPP), switch rates in the real switch task did not differ significantly between groups. Reaction times were significantly longer among PwPP during the real switch task, but this does not appear sufficient to explain our observations of faster switch rates among PwPP, as slower reaction times alone would not be expected to yield a greater number of reported switches. Thus, relatively normal performance in the real switch task among PwPP suggests that their ability to detect changes in local motion direction was not dramatically impaired, in agreement with previous work (Bennett et al., 2016; Carter et al., 2017; Chen et al., 2006).

Instead, our results may suggest that PwPP experienced less stable global SFM percepts, as switch rates during the bi-stable task were significantly faster than for control participants. Models of bi-stable perception suggest that reciprocally connected neural populations with different tuning properties (e.g., selectivity for rightward vs. leftward rotation in depth) may compete when viewing ambiguous bi-stable stimuli, with perception alternating as one population suppresses the other, and then is suppressed in return (Said & Heeger, 2013; Tong et al., 2006; H. R. Wilson, 2003). Our results may suggest that the depth of suppression between competing populations may be weaker among PwPP during SFM perception; weaker suppression might be expected to yield faster switch rates, as the dominant population would be less able to sustain suppression of the competing percept. This would appear consistent with other reports of weaker perceptual suppression in PwPP (e.g., center-surround suppression, temporal masking; da Cruz et al., 2020; Dakin et al., 2005; Herzog & Brand, 2015; Schallmo et al., 2015; Sponheim et al., 2013).

A pair of recent studies examined bi-stable perception among people with schizophrenia using a rotating sphere illusion (Schmack et al., 2015, 2017). Those studies used methods to bias perception towards one rotation direction by stopping and starting the stimulus (continuation bias) or through repeated presentation of unambiguous stimuli that tended to rotate more in one direction (learning bias). Their goal was to examine bi-stable SFM perception under the framework of predictive coding. They found that people with schizophrenia showed less-biased perception in both their continuation and learning paradigms. Although bi-stable switch rates were not reported in these previous studies, our current findings appear generally consistent with their notion of weaker sensory predictions during bi-stable SFM perception in PwPP.

We predicted switch rates would be slower among PwPP compared to controls, based on results in the literature showing slower switch rates during binocular rivalry among people with schizophrenia (Fox, 1965; Miller et al., 2003; Nagamine et al., 2009; Ngo et al., 2011; Xiao et al., 2018; Ye et al., 2019). Contrary to this prediction, we found switch rates in our SFM task were in fact faster among PwPP when compared to healthy controls. There are a number of factors that may explain this apparent discrepancy. Previous studies used a different bi-stable paradigm, binocular rivalry, in which participants are shown two dissimilar stimuli (one in each eye, e.g., orthogonal gratings) that compete for perceptual dominance. This task is thought to particularly engage neural populations selective for low-level image features (e.g., orientation, color), such as those in V1, to drive interocular suppression and perceptual rivalry (Leopold & Logothetis, 1999; Tong & Engel, 2001). Thus, binocular rivalry may depend more strongly on suppression at an earlier stage of neural processing compared to bi-stable SFM. Indeed, a recent study by Cao and colleagues (Cao et al., 2018) showed that switch rates from rotating cylinder and binocular rivalry tasks were only weakly correlated across individuals.

In addition, previous studies have focused specifically on people with schizophrenia *or* bipolar disorder, whereas we examined bi-stable visual perception across the psychosis spectrum. Although we saw significantly faster switch rates among PwPP versus controls overall, and significant differences in the post-hoc comparison between schizophrenia and controls, post-hoc tests between bipolar and control groups were not significant. This may reflect a lack of statistical power due to the smaller sample sizes within these diagnostic sub-groups. Our schizophrenia and bipolar sub-groups showed very similar switch rates to one another, suggesting that bi-stable SFM perception was not dramatically different across different diagnostic categories within the psychosis spectrum.

We hypothesized that abnormal switch rates during bi-stable SFM perception might be related to impaired E/I balance in psychosis. Dysfunction in both excitatory glutamate (Javitt, 2004; Moghaddam & Javitt, 2012) and inhibitory GABA functioning (Gonzalez-Burgos et al., 2010; Volk & Lewis, 2005) have been reported in psychotic disorders. Other studies suggest that bi-stable perception, including in the rotating cylinder illusion, may be linked to E/I balance in visual cortex among healthy adults (Mentch et al., 2019; Robertson et al., 2016; van Loon et al., 2013). Although we observed a relationship between higher inhibitory GABA levels and slower SFM switch rates in a direction that matched previous reports (van Loon et al., 2013), this correlation did not reach statistical significance, and we saw no significant relationships between SFM switch rates and excitatory glutamate or glutamine. Because we saw no significant group differences in GABA levels, nor any significant correlations within groups between GABA and switch rates (after correction for multiple comparisons), we cannot draw any strong conclusions about role of GABAergic inhibition in abnormally fast switch rates among PwPP. A limitation of our study is that MRS data were acquired only in a small region of the mid-occipital lobe, which generally encompasses early visual areas (e.g., V1) with peripheral retinotopic selectivity. Thus, it is not clear to what extent neurochemical concentrations in other brain regions (e.g., those with greater foveal selectivity, and / or higher visual areas such as hMT+) may be linked to perception of bi-stable SFM.

Examining SFM task performance among first-degree relatives allowed us to explore the role of genetic liability for psychosis during bi-stable SFM perception. Across our analyses, results from relatives tended to fall in between controls and PwPP. This pattern of results can be seen both in the average switch rates (Figure 4) and the distribution of percept durations (Figure 3). These results are consistent with the notion of attenuated visual dysfunction among unaffected relatives that falls on a continuum between healthy controls and PwPP (Pokorny et al., 2020), and suggest that faster-than-normal switch rates during bi-stable SFM perception may be linked in part to genetic liability for psychosis.

## Data Availability

All data collected as part of the Psychosis Human Connectome Project and will be made publicaly available later this year or upon reasonable request to the authors.

## Acknowledgments

This work was supported by funding from the National Institutes of Health (U01 MH108150). Salary support for MPS was provided in part by K01 MH120278. Salary support for KWK was provided in part by T32DA037183. Support for MR scanning at the University of Minnesota Center for Magnetic Resonance Research was provided by P41 EB015894 and P30 NS076408. This work used tools from the University of Minnesota Clinical and Translational Science Institute that were supported by UL1 TR002494.

We would like to thank and acknowledge Kimberly B. Weldon, Li Shen Chong, Marisa J. Sanchez, Rohit S. Kamath, Samantha A. Montoya, Victoria Espensen-Sturges for contributing to data collection and management, as well as Caroline Demro for help with data processing, and Cheryl A. Olman for supporting the study design.

## Supplemental figures

**Supplemental Figure 1.**
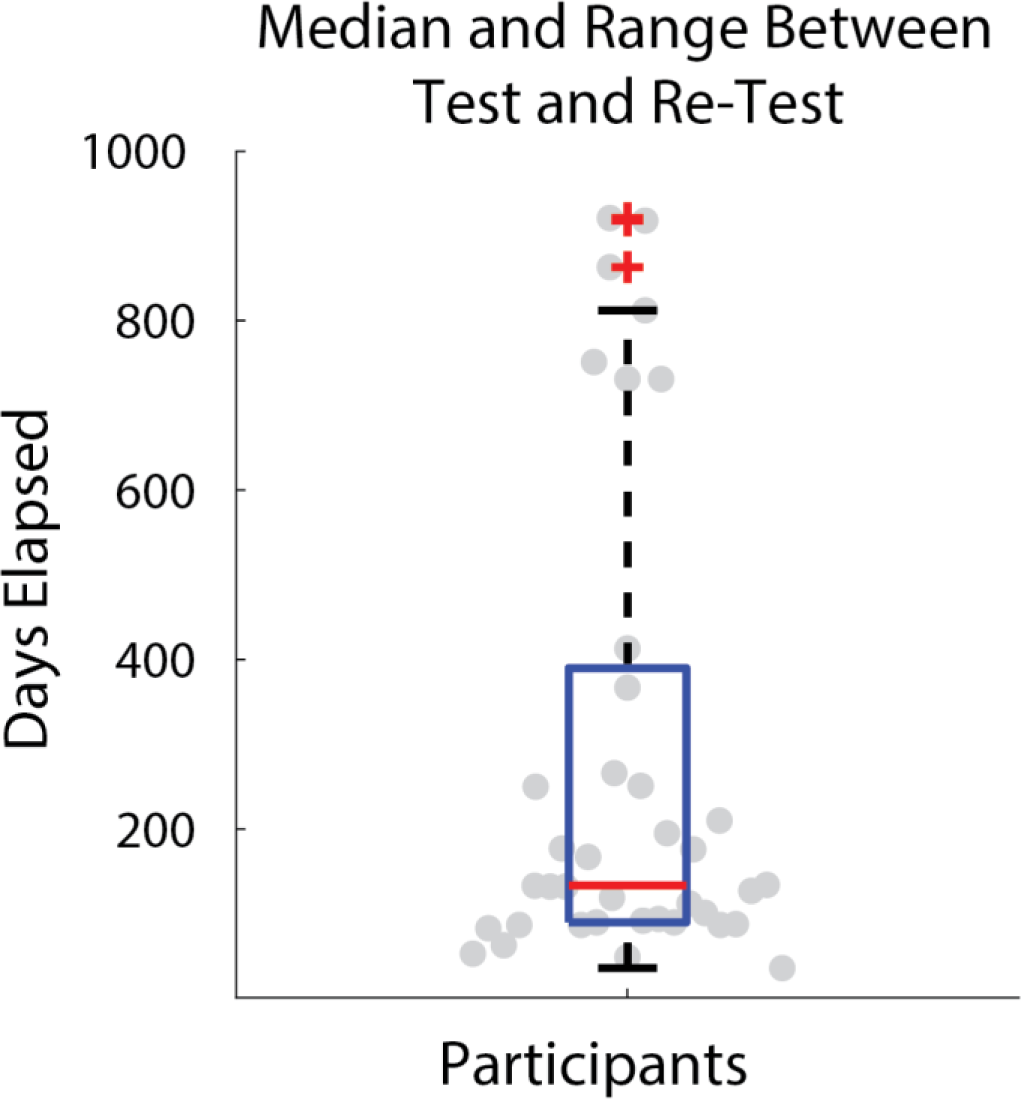
Median and range of days between test and re-test sessions. Box plot of 1.5 x the interquartile range (dashed line), 25% - 75% quartiles (blue), and median (red) of the number of days between the first and second sessions for all participants, who took part in both sessions. Outliers are shown with red crosses.

**Supplemental Figure 2.**
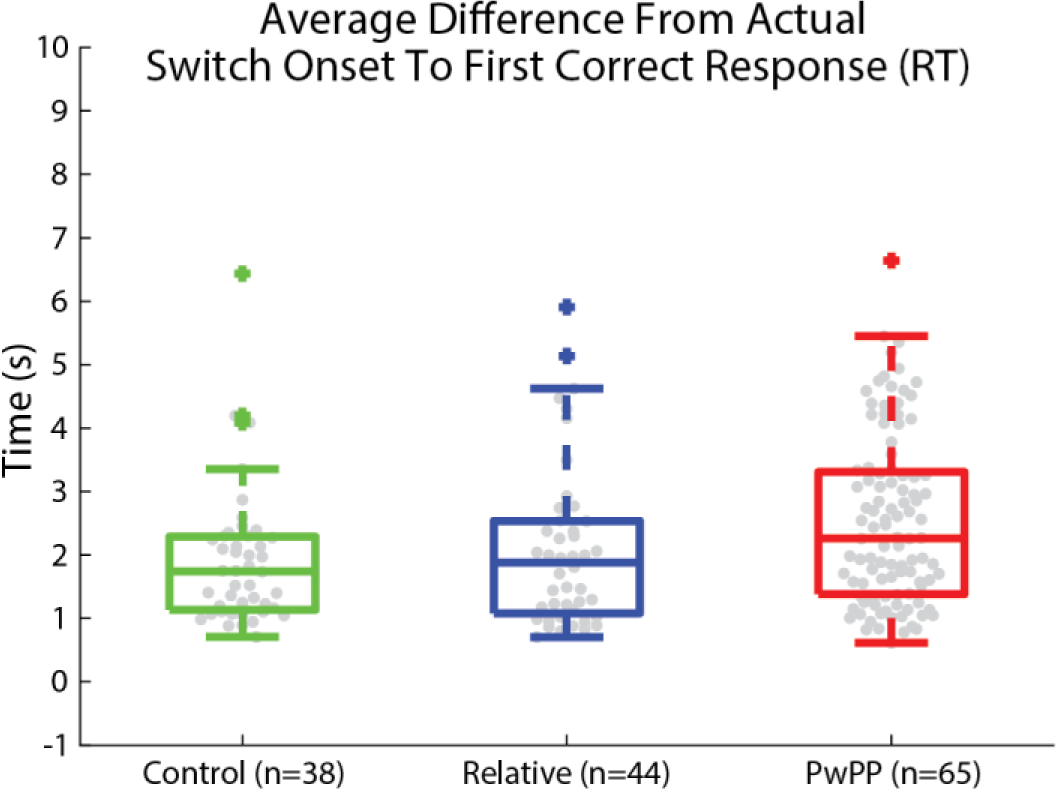
Average difference across groups between the physical switch onset and the first correct response. Boxplots show median (middle line) 25-75% (box), 1.5 x the interquartile range (whiskers), and outliers (pluses), as well as individual data points (gray).

**Supplemental Figure 3.**
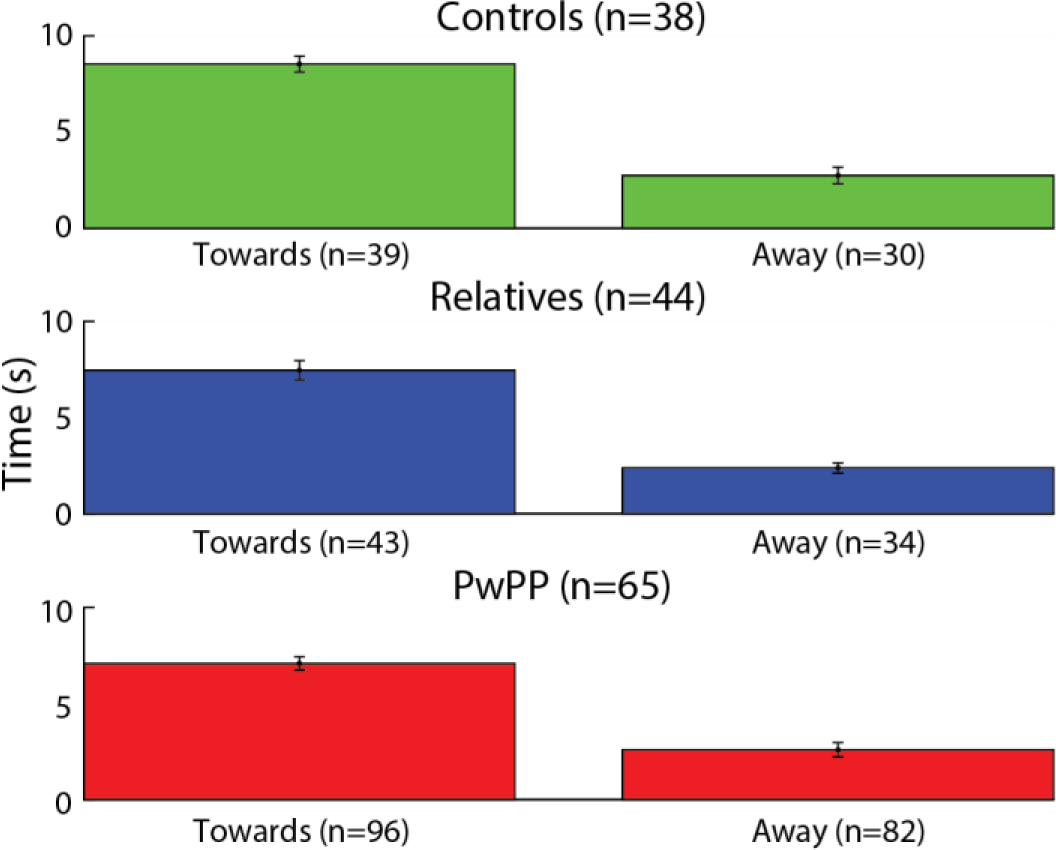
Average percept duration for responses made toward or away from the direction of the physical switch. Plotted is the average time between reporting a switch and reporting the next switch (either away from or towards the actual physical rotation direction), for each group. Error bars represent the standard error of the mean.

**Supplemental Figure 4.**
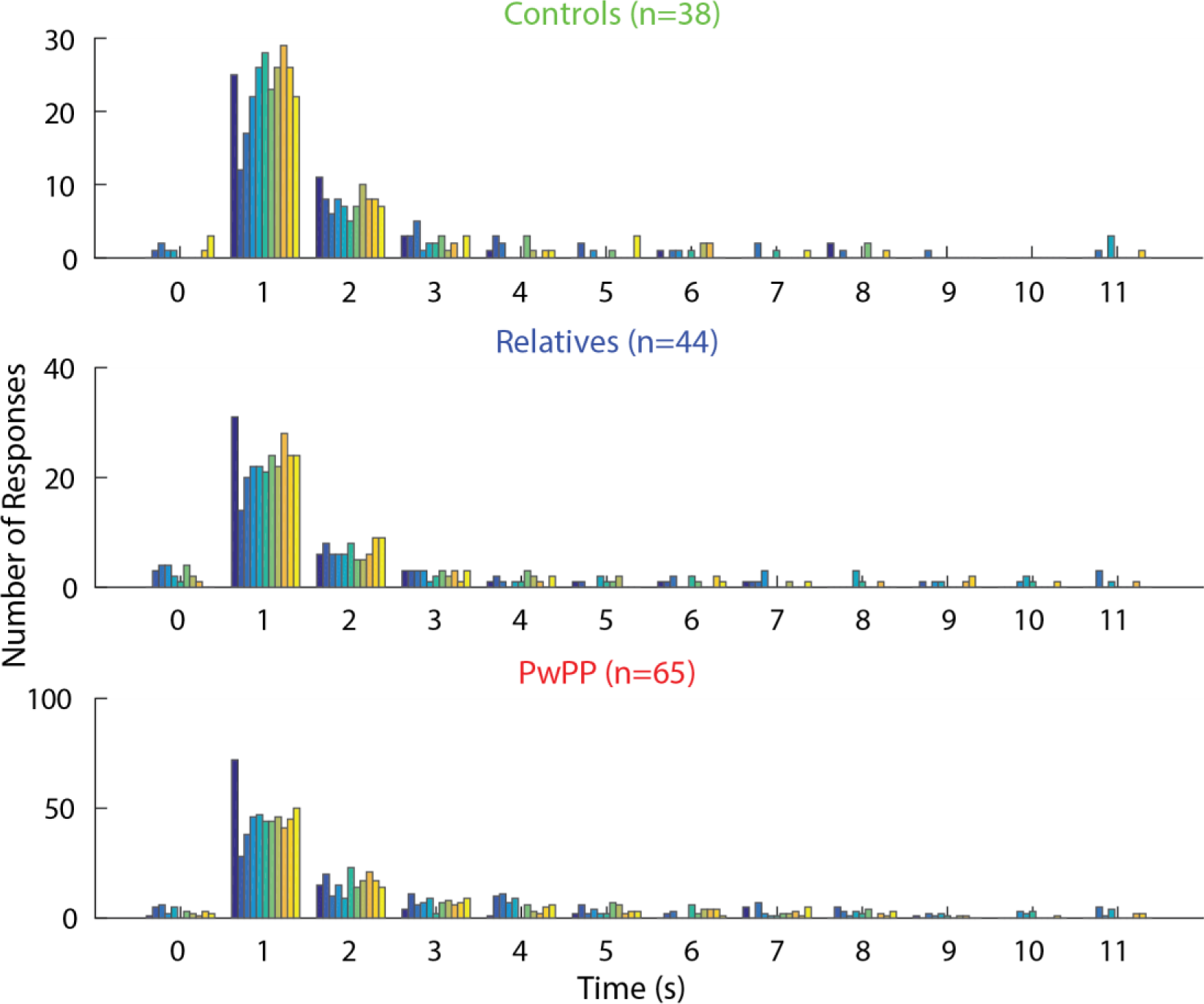
Distribution of total responses made by response time. Plotted are the number of responses (y-axis) made within each time bin (x-axis) for 11 s following a stimulus direction change. Colored bars represent physical switch number (11 total physical switches per subject) ordered from first to last presented switch.

**Supplemental Figure 5.**
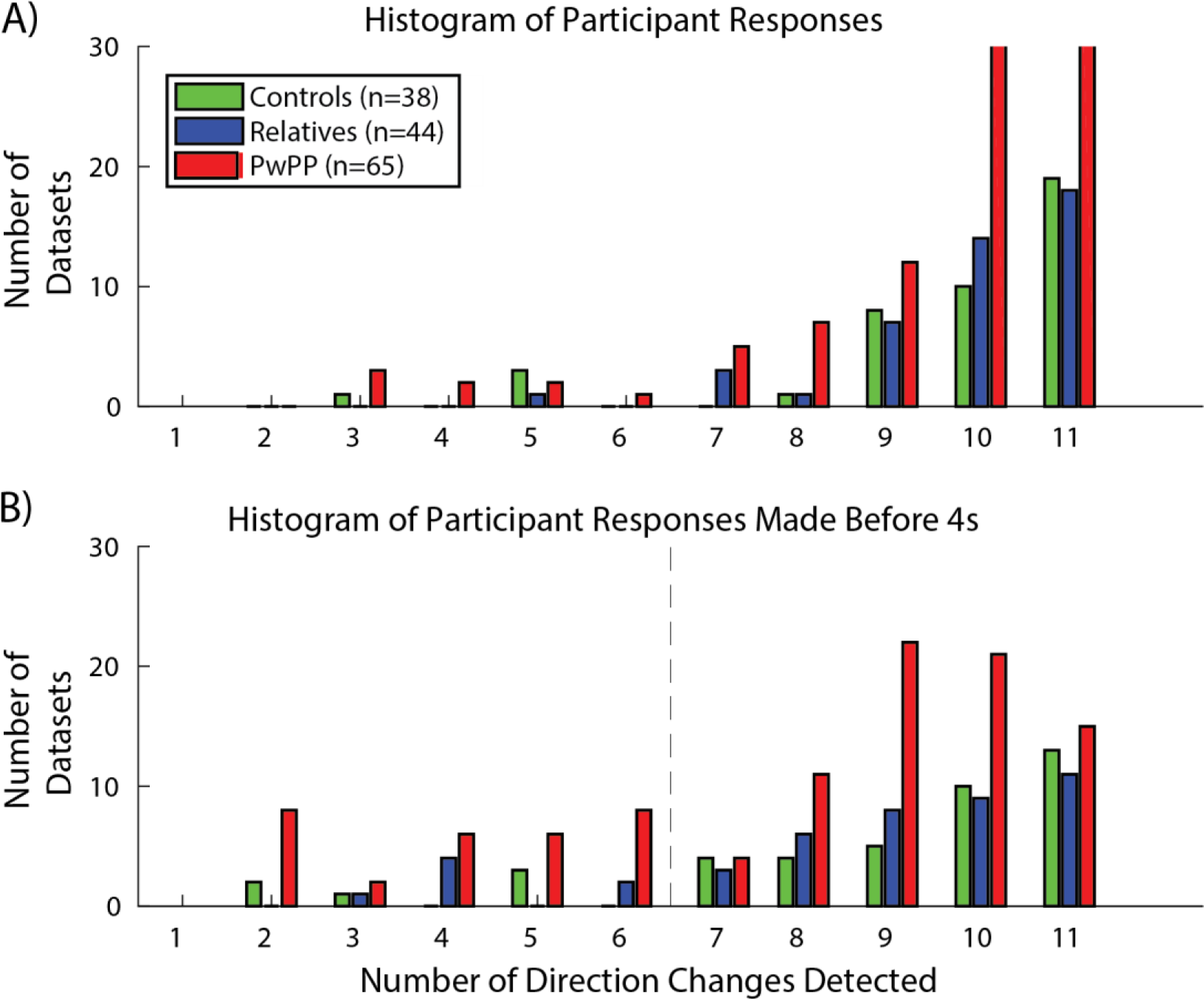
Distribution of the number of responses made across all participants. Plotted in panel A are the number of datasets where participants responded correctly to the corresponding number of physical switches. Plotted in panel B are the same data thresholded by a response time of 4 seconds. The colored bars represent the three groups and the vertical dashed line represents a cutoff of 63.4% or that the participant responded to at least 7/11 physical switches.

**Supplemental Figure 6.**
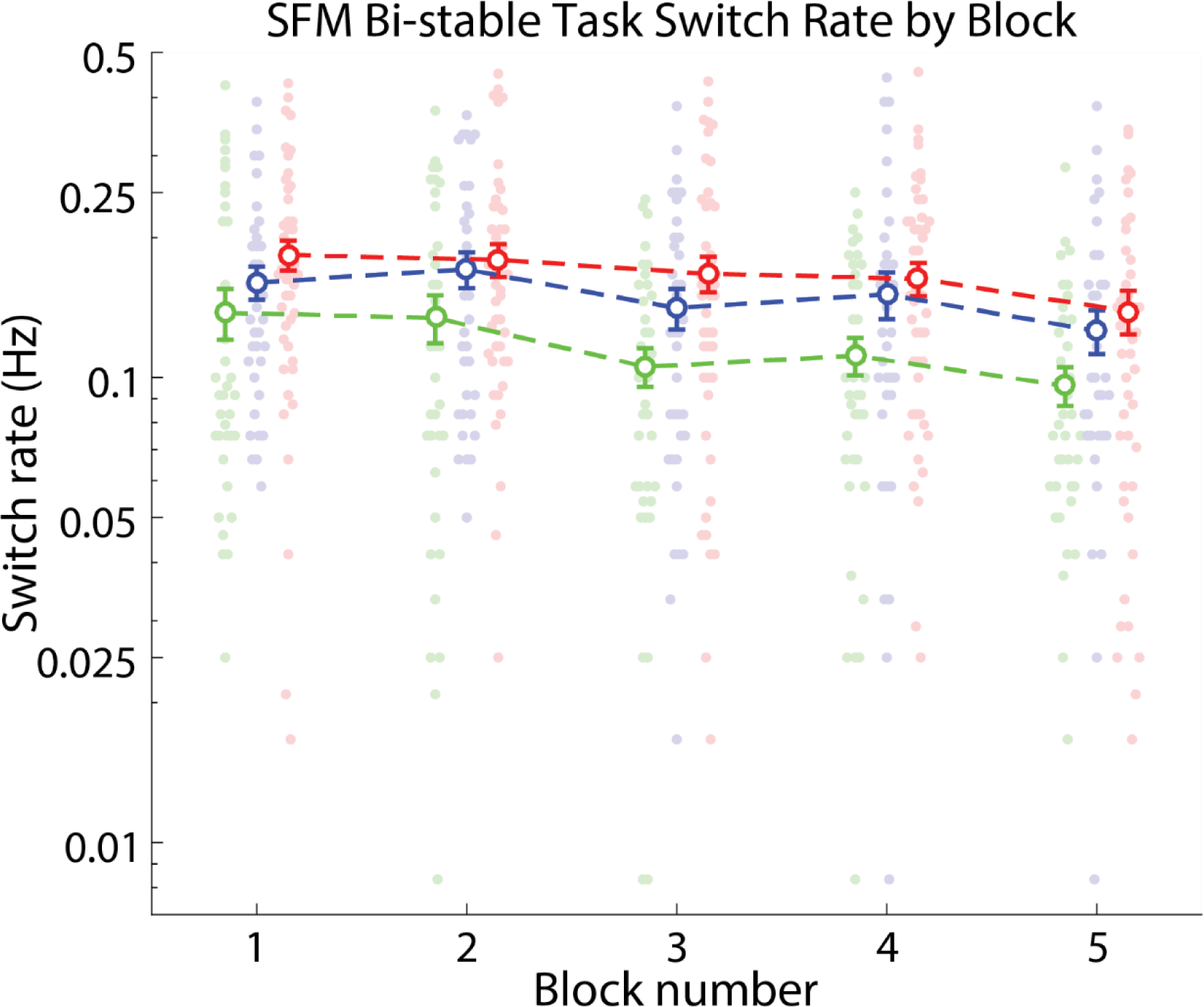
Switch rate for each group over time (block). Plotted are the average switch rates during each block for all three groups (controls (n=37) – green, relatives (n=37) – blue, PwPP (n=48) – red). Error bars represent the standard error of the mean.

**Supplemental Figure 7.**
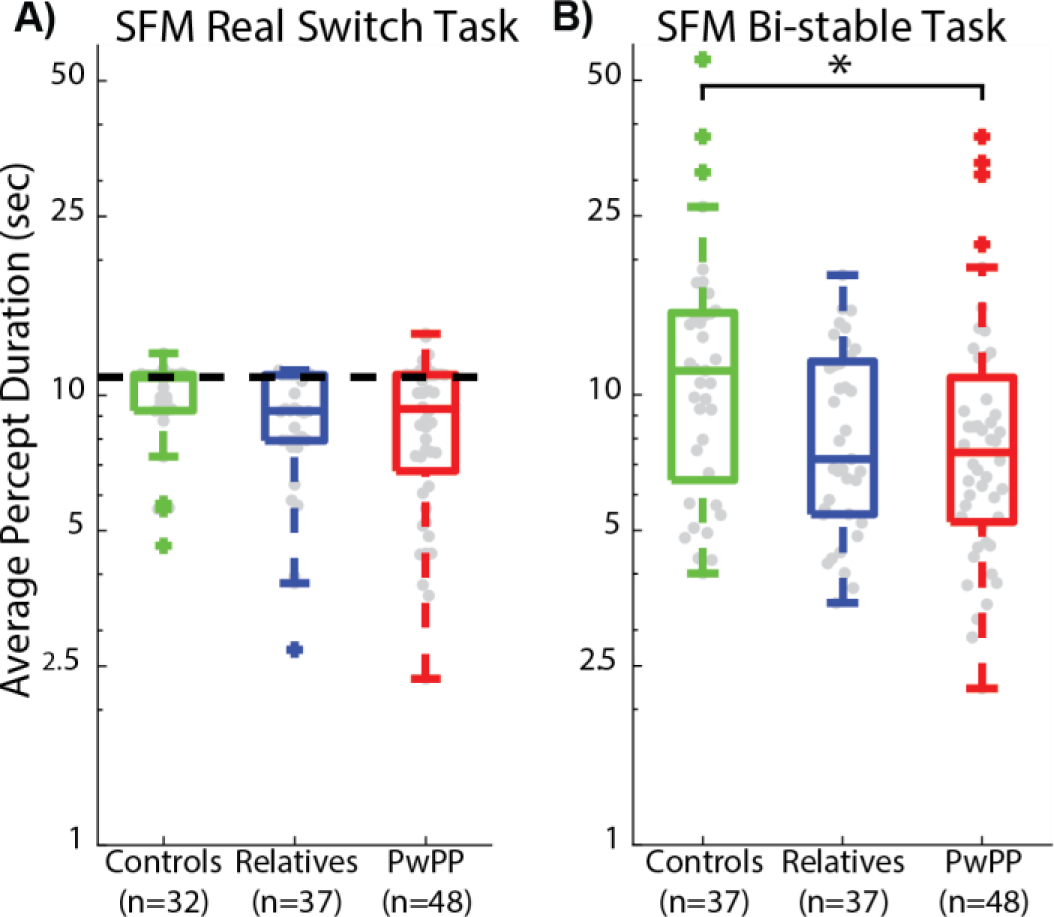
Average percept duration for the three groups in (A) the real switch and (B) bi-stable tasks. Shown are box plots displaying median, 25%-75% quartiles, and 1.5 x the interquartile range for the three groups (controls – green, relatives – blue, PwPP – red). Outliers are shown overlaid on crosses. Dashed line in A shows average percept duration for physical stimulus changes in the real switch task.

**Supplemental Figure 8.**
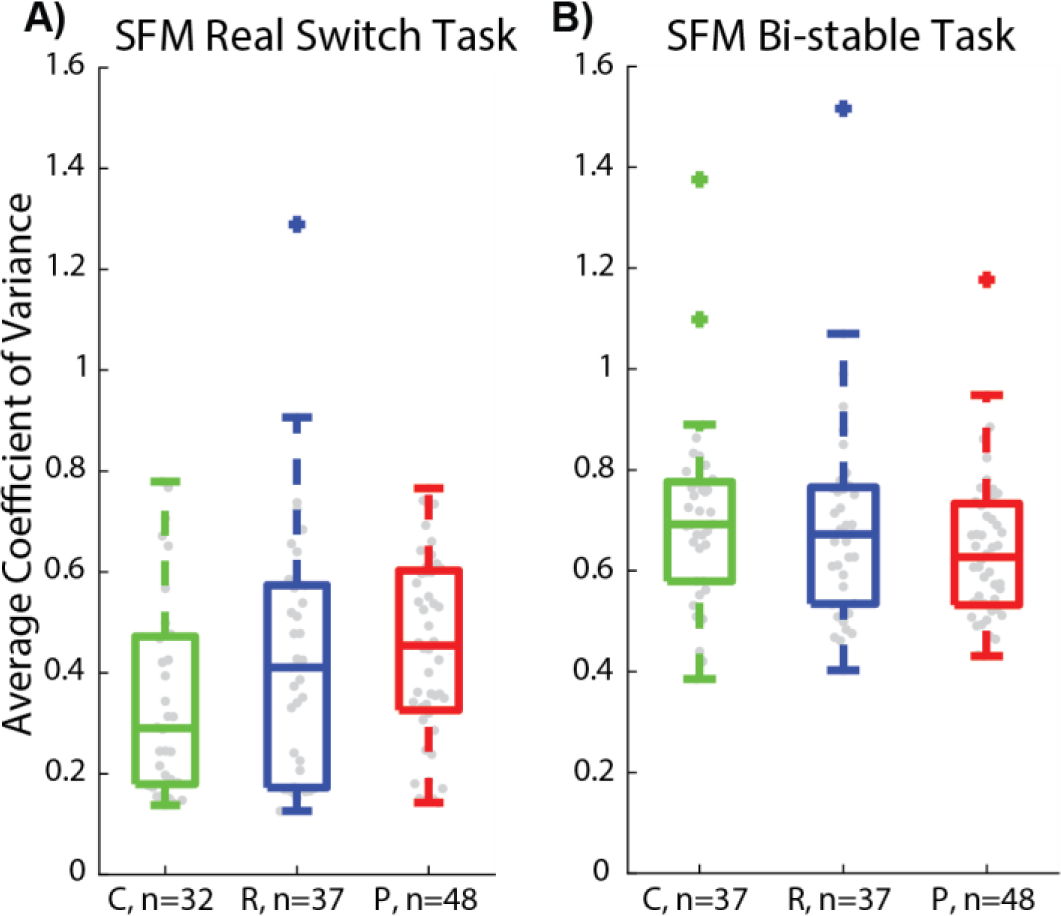
Average coefficient of variance (*SD* divided by the mean) for percept durations in both the (A) real switch and (B) bi-stable tasks. Shown are box plots displaying median, 25%-75% quartiles, and 1.5 x the interquartile range for the three groups (controls – green, relatives – blue, PwPP – red). Outliers are shown overlaid on crosses.

**Supplemental Figure 9.**
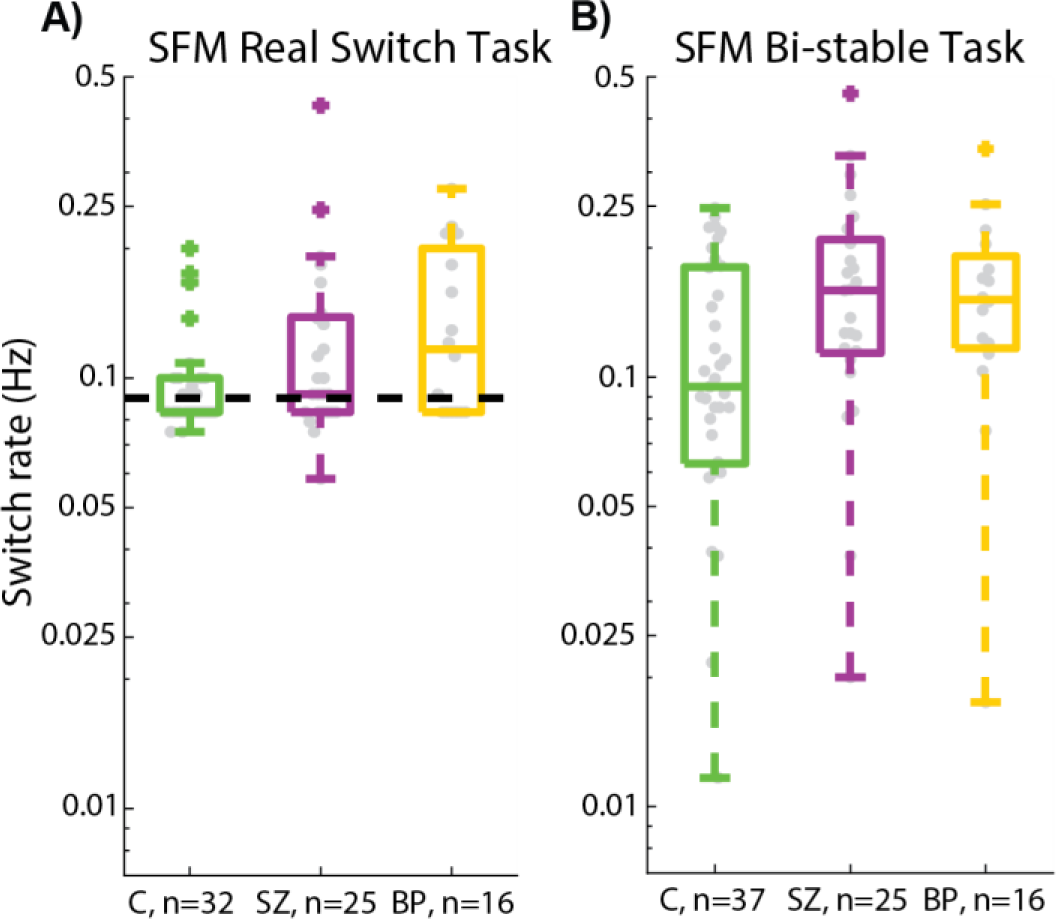
Average switch rate for the control, bipolar, and schizophrenia groups in (A) the real switch and (B) bi-stable tasks. Shown are box plots displaying median, 25%-75% quartiles, and 1.5 x the interquartile range for the three groups (controls (n=37) – green, schizophrenia (n=25) – purple, bipolar (n=16) – yellow). Outliers are shown overlaid on crosses.

**Supplemental Figure 10.**
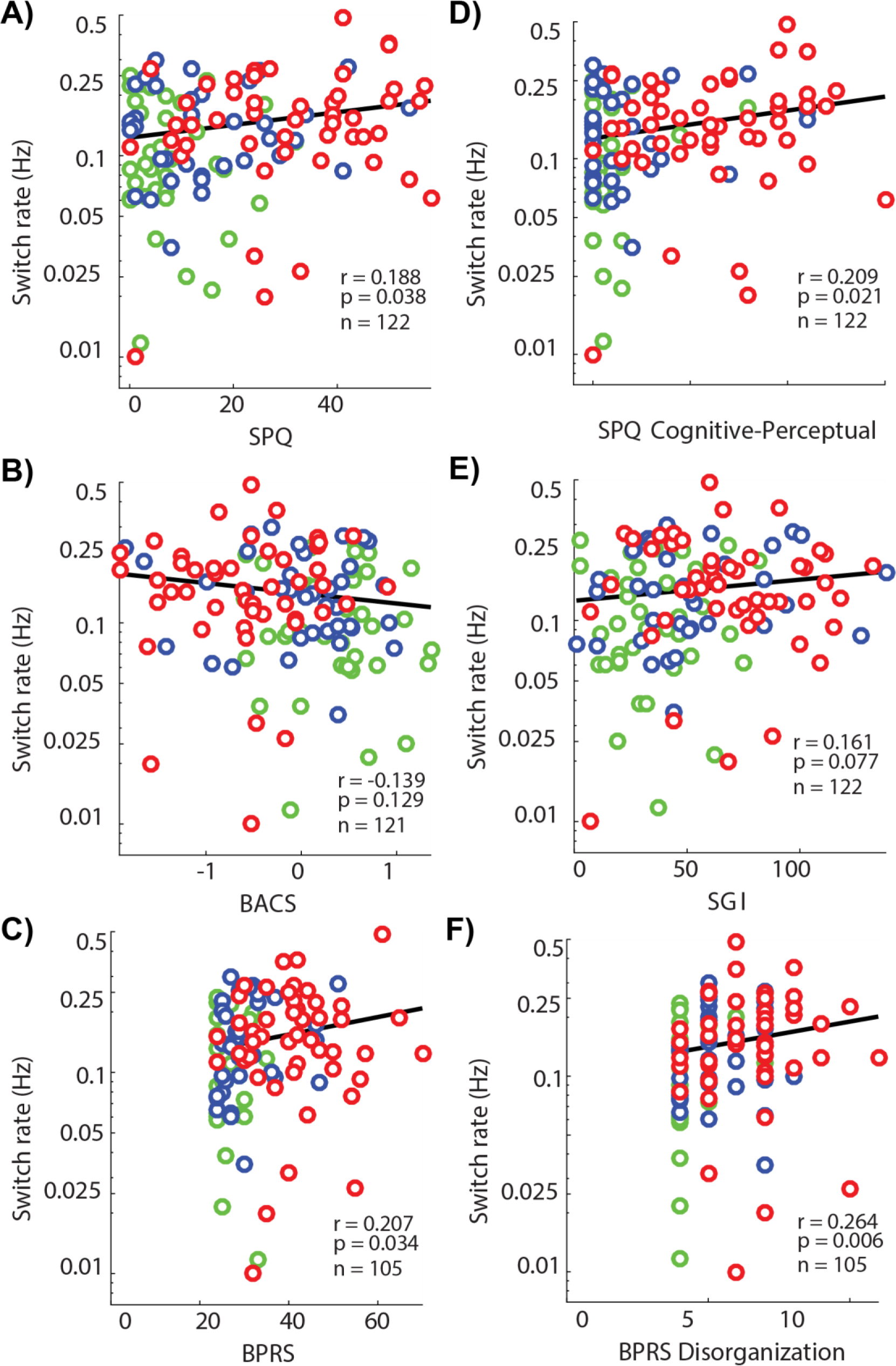
Bi-stable switch rate correlations with measures of psychotic symptomology. Relationship between switch rate and 6 measures of psychotic symptomology: (A) SPQ, (B) BACS, (C) BPRS, (D) SPQ Cognitive-Perceptual Factor, (E) SGI, (F) BPRS Disorganization Factor. Plotted points represent individual participants broken down by color for different groups (controls (n-31) – green, relatives (n-37) – blue, PwPP (n=48) – red). Y-axes show bi-stable switch rates (Hz) and x-axes show clinical test scores. R-values indicate Spearman correlation with retest data excluded. P-values are not corrected for multiple comparisons.

**Supplemental Figure 11.**
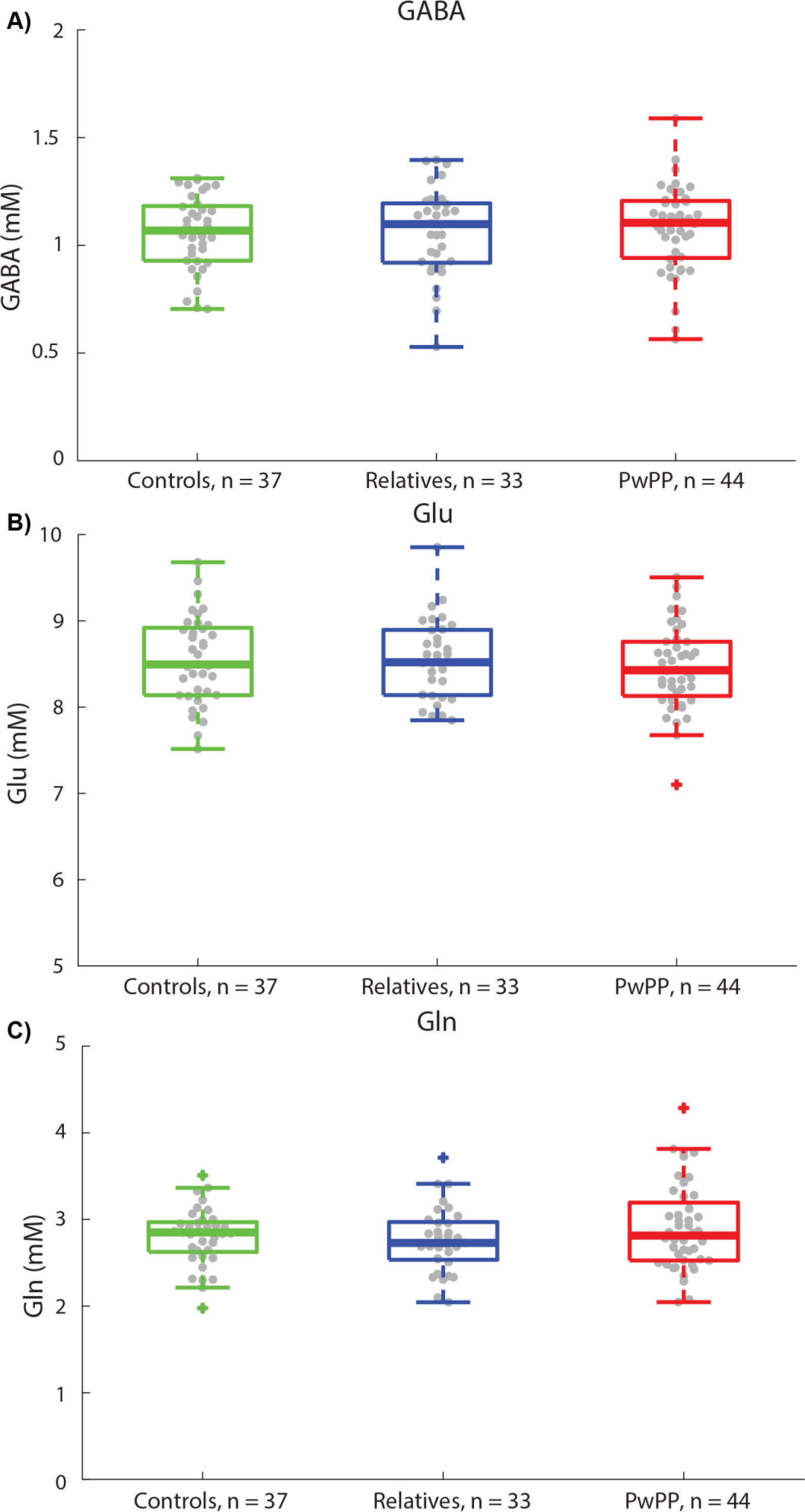
MRS metabolite concentrations in the three groups. Average metabolite concentration (mM) for the three selected neurochemicals: (A) GABA, (B) glutamate (Glu) (B), and (C) glutamine (Gln). Shown are box plots displaying median, 25%-75% quartiles, and 1.5 x the interquartile range for the three groups (controls – green, relatives – blue, PwPP– red). Outliers are shown overlaid on crosses.

**Supplemental Figure 12.**
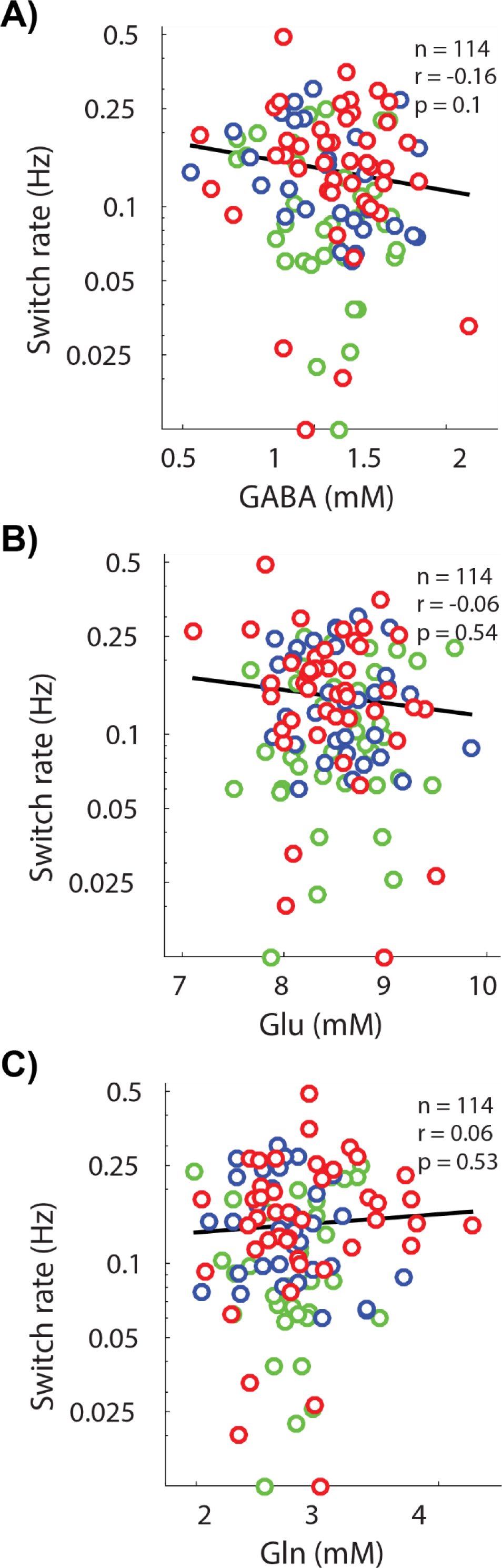
Bi-stable switch rate correlations with neurochemical concentrations. Correlations between switch rate (Hz) and concentrations of three metabolites: (A) GABA, (B) glutamate, (C) and glutamine. Plotted on the y-axis are bi-stable switch rates (Hz) and on the x-axis neurochemical concentrations (mM). Plotted points represent individual participants broken down by color for different groups (controls (n=37) – green, relatives (n=33) – blue, PwPP (n=44) – red)

